# What do Australians affected by cancer think about oncology researchers sharing research data: a cross-sectional survey

**DOI:** 10.1101/2023.05.01.23289334

**Authors:** Daniel G. Hamilton, Sarah Everitt, Matthew J. Page, Fiona Fidler

## Abstract

**Objectives:** To characterise the attitudes of Australians affected by cancer towards the sharing of de-identified research data with third parties, including the public.

**Design, setting, participants:** Anonymous online survey between October 2022 and February 2023 of adult Australians previously diagnosed with cancer.

**Main outcome measures:** Self-reported attitudes towards the sharing of human and non-human data, and the hypothetical sharing of their anonymised medical information and responses to the survey.

**Results:** 551 respondents contributed data to the survey. There was strong support for cancer researchers sharing non-human and de-identified human research data with medical doctors (90% and 95% respectively) and non-profit researchers (both 94%). However, this declined when participants were asked whether data should be shared with for-profit researchers (both 64%) or posted publicly (both 61%). When asked if they would hypothetically consent to researchers at their treatment location collecting and sharing their de-identified data publicly, only half agreed (50%). In contrast, after being shown a visual representation of the de-identified survey data, 80% of respondents supported sharing it publicly. A further 10% also supported public sharing of some of the survey data, with the most frequently desired information to be withheld including education history and levels of trust in healthcare stakeholders.

**Conclusions:** Australians affected by cancer support the sharing of research data, particularly with clinician and non-profit researchers. Visualisation of the data to be shared may also enhance support for making research data publicly available. These results should help alleviate any concerns about research participants’ attitudes on data sharing, as well as boost researchers’ motivation for sharing.

---

The last three years have seen an increase in discussions about data sharing and Open Science more broadly in Australia. On one side, we saw the first steps towards a national Open Access Strategy, as well as Australia’s adoption of the United Nations Educational, Scientific and Cultural Organization’s (UNESCO) international framework on open science in 2021 [1]. Australian funding agencies have also been put under pressure to strengthen their public access policies as nations like the United States instruct federal funding agencies to make publications and their supporting data publicly accessible without an embargo or cost. The National Health and Medical Research Council (NHMRC), which directed $1.6 billion to cancer-related research between 2013-2021 and is a signatory of the San Francisco Declaration on Research Assessment (DORA), is among them [2]. However, in contrast, the same period bore witness to several well-publicised and serious leaks of Australians’ health information from private companies (e.g., Medibank & AHM, Medlab Pathology, Australian Clinical Labs), public healthcare providers (e.g., Ambulance Tasmania) and contractors working with government departments (e.g., PNORS Technology Group). Combined, these incidents exposed tens of millions of Australians’ data. Consequently, this may have eroded previously high levels of support among the general Australian public and people living with cancer for medical researchers collecting and sharing health data observed [3,4].

Greater availability of the data underpinning published research comes with many benefits to scientists and society [5,6]. However, data sharing in oncology is not yet the norm [7,8], which has impeded attempts to replicate cancer research [5]. Commonly cited challenges to sharing data include legislative and regulatory barriers, time and resource burdens and concerns about negative impacts on academic performance (e.g., loss of publishing and funding opportunities) [9]. Consequently, several authors have explored the attitudes of medical research stakeholders towards improving access to researchers’ data [9,10]. However, this body of research has largely focussed on the attitudes of researchers, institutions, funders, and publishers. Less attention has been paid to what the key contributors to, and ultimate consumers of medical research – patients – think of these practices [11,12]. Furthermore, research on this topic has focussed on certain data types (e.g., omics data [13,14]), study designs for which public sharing of data is not routine (e.g., clinical trials [12,15,16]) or specific cancer types (e.g., breast cancer [4]). Therefore, this study aims to contextualise and build upon previous research into public opinions on data sharing by engaging Australians affected by cancer and characterising their views on the sharing of de-identified research data with third parties, including the public.

## METHODS

A short, cross-sectional survey designed to capture the views on data sharing of Australians affected by cancer was opened on October 27th, 2022. Any person over the age of 18, who was an Australian citizen or resident, able to comprehend English, and had been previously diagnosed with a cancer of any kind was eligible to participate. We summarise key aspects of the methods below. Further information on the study methods is included in Appendix 1 and on the project’s Open Science Framework page [17]. The findings of this study are reported in accordance with the Checklist for Reporting of Survey Studies (CROSS) guidelines (Appendix 1) [18].

### Survey design

The survey contained 23 questions separated into five sections (Appendix 2). The first section collected demographic information from participants, as well as their trust levels in key Australian healthcare stakeholders. The second section explained the concept of ‘research data’ and the commonly discussed reasons for and against sharing, with the order of reasons for and against sharing being randomised for each participant. The third checked participants’ comprehension of our definition of research data using real-world data from a study investigating viral and fungal infections in people diagnosed with lymphoma [19], as well as captured whether participants were familiar with these concepts. The fourth characterised participants’ general views on both the sharing of human and non-human research data, and the hypothetical sharing of de-identified research data containing their medical information with four different recipients (medical doctors, non-profit researchers, for-profit researchers, and the public). The fifth and final section provided a visual representation of what the de-identified survey data would look like, incorporating the participant’s responses into one of the rows (Figure 1), then proceeded to ask the participant for their views on: i) whether and how long the data should be retained, ii) whether they would hypothetically allow the research team to share it publicly, and iii) if not, which data, if any, they wished to be withheld and the reasons why.

**Figure 1.**
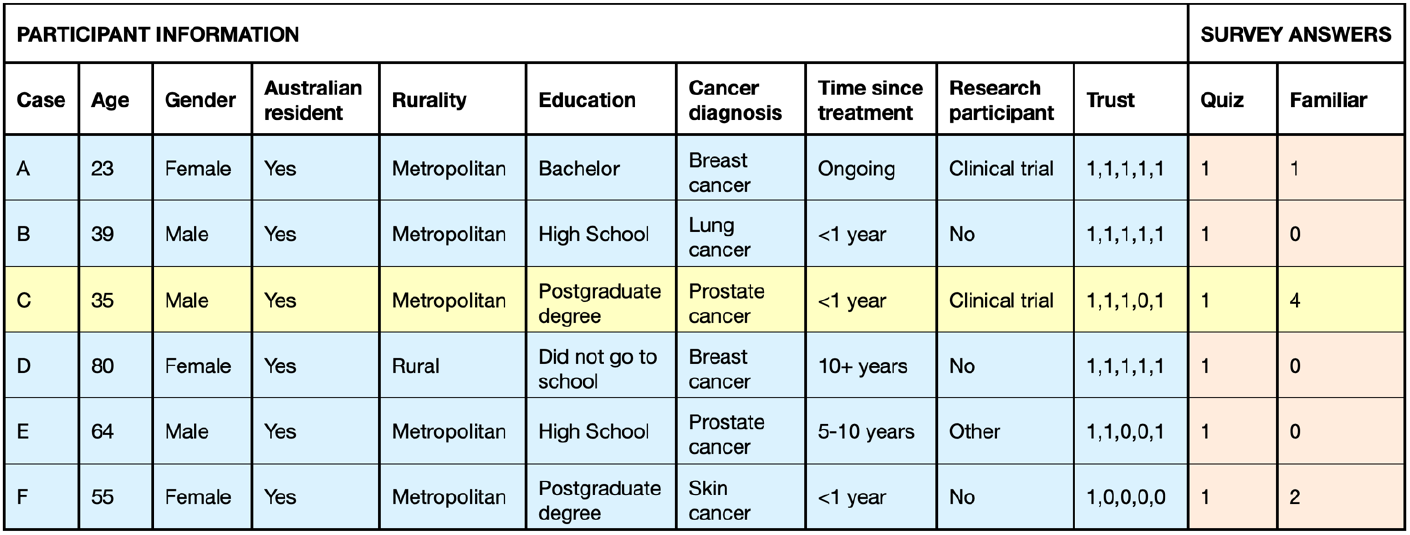
Visual aid used to display what the research data from the survey would look like, with the data contributed by each participant displayed in the yellow row. (Note: The data in the yellow row below has been created for instructional purposes.)

### Survey piloting and distribution

The survey was pilot tested between August and September 2022 by ten individuals of varying backgrounds (Appendix 1). Following this, the readability of the survey transcript was then assessed using the SMOG (Simple Measure of Gobbledygook) scale [20], and further language edits were made to ensure that the average survey readability score stayed at a seventh-grade reading level. Qualtrics Solutions’ Online Survey Software (Qualtrics, Provo, UT) was used to host the survey and three passive recruitment strategies were used to advertise it (social media, cancer organisation newsletters and physical flyers).

Several features were also incorporated into the survey to prevent data entry errors and identify submission of fraudulent responses.

### Statistical analysis

Continuous data are presented as medians and interquartile ranges (IQR) and categorical data are presented as counts and proportions. Risk differences (RD) and associated 95% confidence intervals (CI) were calculated to assess differences in support for data sharing according to i) human versus non-human data, ii) trust in multiple stakeholders, iii) previous participation in research, iv) comprehension of, and v) familiarity with data sharing, and vi) the order in which respondents were shown the reasons for and against sharing. Deductive content analysis was used to analyse qualitative data, whereby emergent themes were classified using pre-established coding criteria [11]. The frequency with which themes occurred in responses were then explored. Quantitative analyses were performed in R (R Foundation for Statistical Computing, Vienna, Austria, v4.2.1) and qualitative analyses were performed in Microsoft Excel.

### Ethics approval

The study protocol was reviewed and approved by the University of Melbourne’s STEMM2 Human Ethics Committee (Project ID: 2022-22111-32090-5) prior to survey piloting and recruitment.

## RESULTS

### Survey participants

Between the survey activation and closure dates, 736 people entered the survey, of which 595 consented to participate, 584 provided at least one answer to the survey, and 551 passed all fraud checks. The median completion time was 12 minutes (IQR: 9-16 minutes). Table 1 shows the characteristics of the sample. The median age of participants was 66 years (IQR: 59-73) and 84% identified as female (N=457). Most participants indicated that they live in a metropolitan area (60%, N=330), followed by small and large rural towns (29%, N=158), then remote and very remote communities (8%, N=45) and were very evenly distributed across the top four levels of education, with a small minority having only completed primary school (1%, N=5). Participants reported being affected by 34 different cancer types, with the most reported diagnosis being breast cancer (54%, N=292), melanoma of the skin (6%, N=30), prostate cancer (5%, N=28), lung cancer (5%, N=29), colorectal cancer (4%, N=23) and lymphoma (3%, N=16). More than a third of participants reported that they had previously participated in a clinical trial (9%, N=49) or non-trial health-related research project (29%, N=156).

**Table 1.**
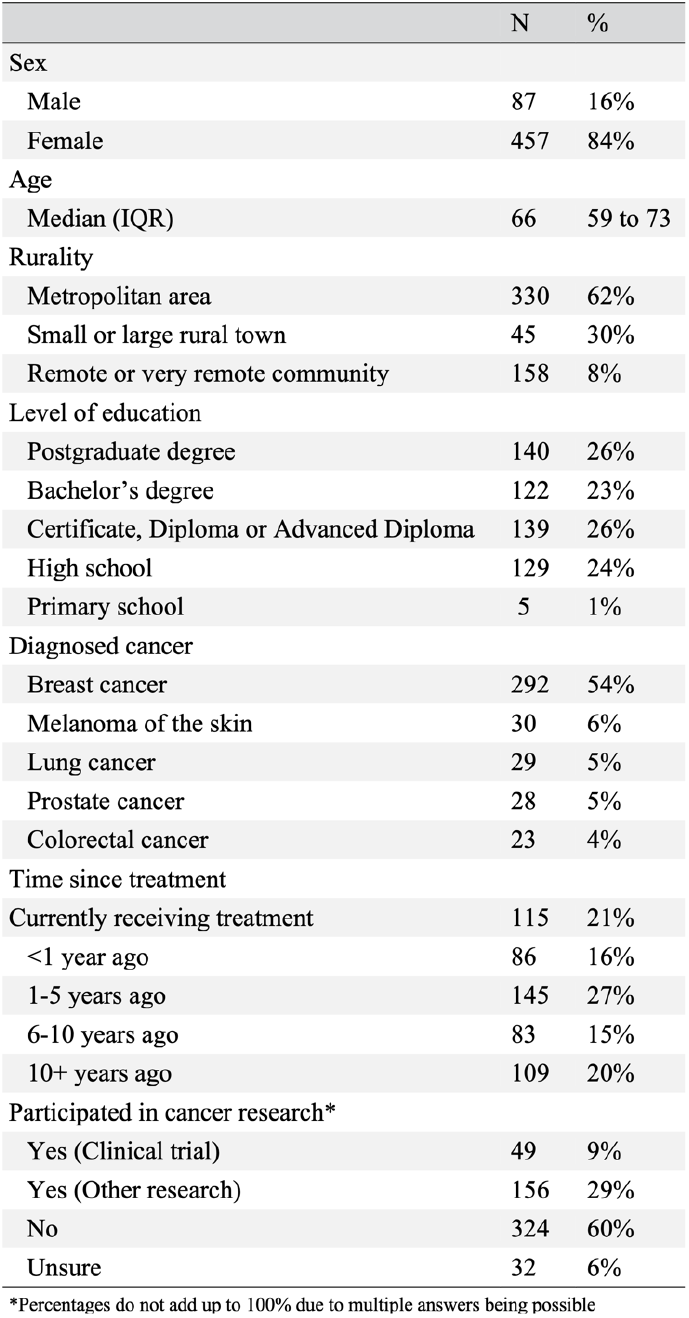
Characteristics of the sample.

### Trust in healthcare stakeholders

Many participants reported trusting their General Practitioner (96%, N=507), other medical doctors in Australia (84%, N=430) and researchers at Australian universities (81%, N=414) with their personal information (Figure 2). However, respondents were less trusting of researchers at Australian companies (34%, N=169) and the Australian government (45%, N=224). Of the participants that answered at least two questions, 92% (N=477) reported trusting multiple groups with their personal information.

**Figure 2.**
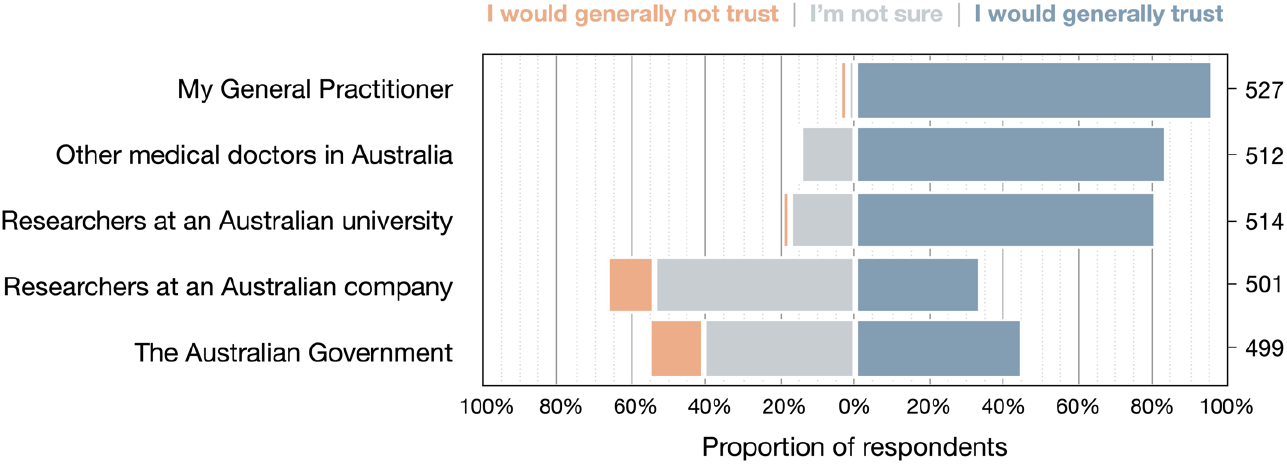
Levels of trust in key clinical medical and research stakeholders.

### Knowledge and general views on data sharing

Most respondents answered the comprehension check correctly (82%, N=347). Similarly, 82% of respondents reported that they had either heard of (15%, N=69), knew a little (53%, N=242), a lot (13%, N=60), or were experts (1%, N=5) on the topic of data sharing. The most common reasons for respondents’ familiarity with data sharing included: general interest in science (51%, N=188), previous participation in health research (25%, N=92) and/or working in health (19%, N=70). When asked to estimate what percentage of cancer research currently shares research data, the median guess was 40% (IQR: 20-60%).

Of the respondents who passed the comprehension check, most reported that they felt cancer researchers should regularly share data derived from human and non-human participants with all four recipients (Figure 3). There was particularly strong support behind the sharing of both data types with medical doctors and non-profit researchers. Interestingly, attitudes towards sharing data with these four groups were similar regardless of whether the data were derived from human or non-human participants.

**Figure 3.**
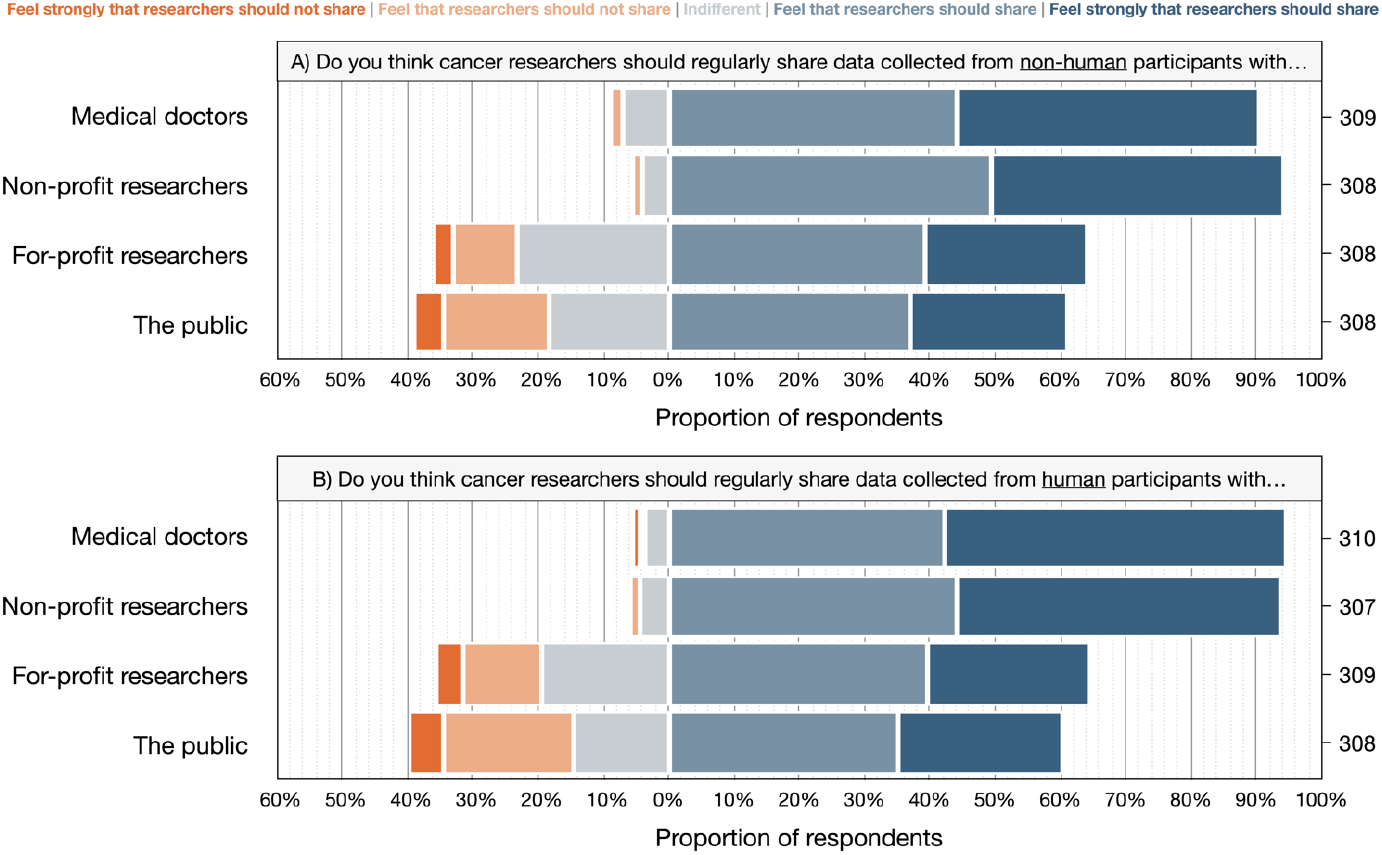
Respondents’ views on the general sharing of research data derived from non-human (A) and human (B) research participants.

### Views on the sharing of their data

When asked hypothetically if they would consent to medical researchers collecting their medical information (e.g., blood pressure measurements, pathology results, prescribed treatments) for a research project, and sharing de-identified data with the same four recipients, most respondents indicated that they would consent to researchers sharing data with medical doctors (99%, N=303) and non-profit researchers (95%, N=291) (Figure 4). However, fewer respondents were supportive of researchers sharing de-identified data with for-profit researchers (56%, N=171) or the public (50%, N=152).

**Figure 4.**
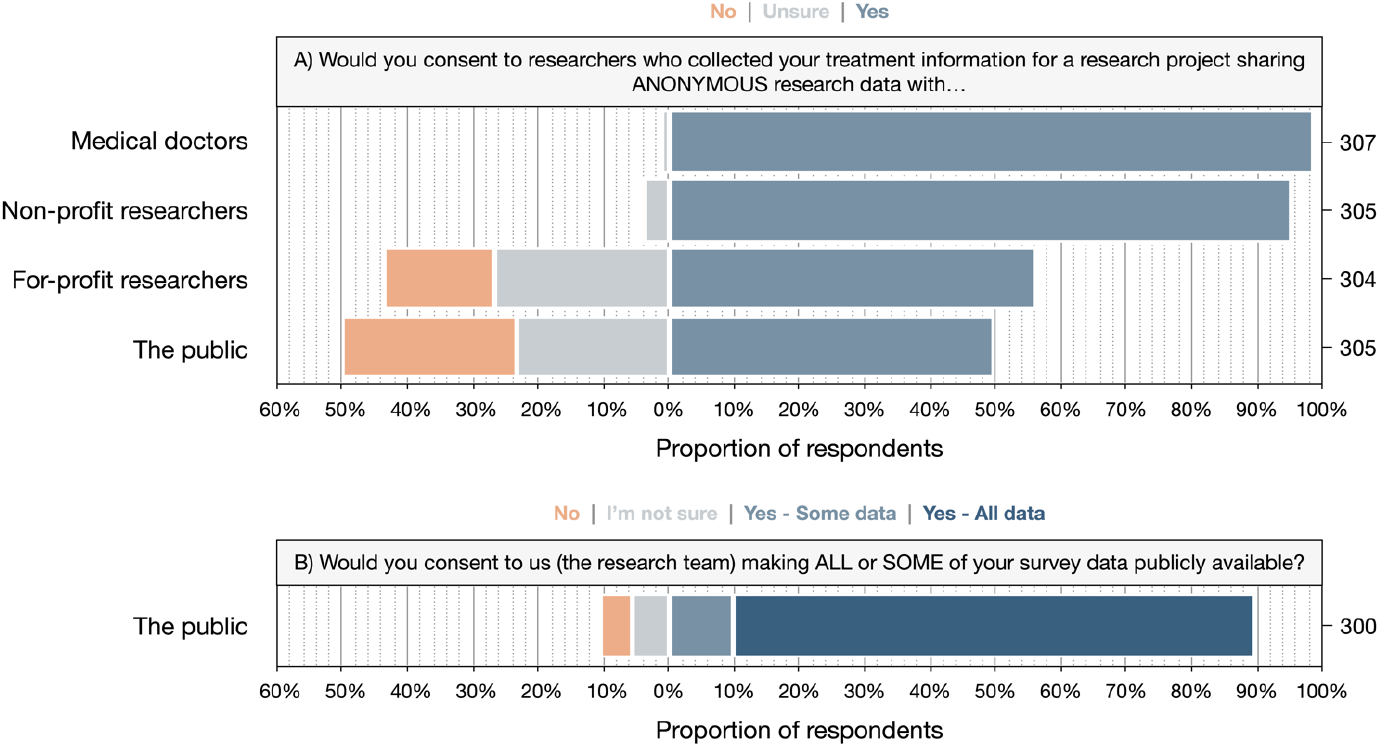
Respondents’ views on the hypothetical sharing of de-identified data containing their treatment information (A) and responses contributed to the survey (B).

In contrast, when participants were shown a visualisation of the survey’s de-identified data incorporating responses they had given (Figure 1), and asked hypothetically whether they would consent to us posting de-identified survey data publicly, 80% (N=240) indicated that they would consent to posting all data publicly, 10% (N=29) reported that they would consent to some data being posted publicly, and 10% (N=31) stated they would not consent or were uncertain. Almost three-quarters of respondents also added that they did not support permanent deletion of this survey’s data following publication of the results (71%, N=215) and were supportive of the research team retaining the data indefinitely (73%, N=221).

Of the 29 that reported that they would only agree to sharing some data, the data respondents most frequently wished to be withheld were the answers to the education (52%, N=15), comprehension (31%, N=9), trust and familiarity (each 24%, N=7) questions. Furthermore, when the 60 respondents who did not support full public release of the survey data were asked what their concerns would be, the most commonly mentioned themes related to re-identification concerns (N=14), misuse by subsequent users of the data (N=12), loss of control of their information (N=10), then potential repercussions for the respondent or other participants (N=2). Several illustrative statements that touched upon these themes are reported in Table 2.

**Table 2.** Example concerns participants had with regards to the hypothetical public posting of the survey data.

| Theme | Subtheme | Example quote |
| --- | --- | --- |
| Confidentiality concerns | Protecting anonymity | "Any possibility of identification, illegal access to my personal data and future identification of me." |
|  | Reassurance | "... [However], I cannot see any personal data that could be traced back to me so I would be comfortable about sharing this survey data." |
| Misuse of data | Commercial research | "Private enterprise using [the data] for profit" |
|  | Misinterpretation | "...I'm concerned how [secondary users] will interpret the data, and what misinformation may then be created." |
|  | Malfeasance | "That [data] would not be used for the right reasons, e.g., for finding a cure for cancer." |
| Loss of control over information | Data recipients | "No control over the purpose for which that data is used, how it is stored" |
|  | Data generators | "Overall, no qualms as long as the public could freely access the data and not have it the published data behind a paywall - especially if the research was government funded." |
|  | Data security | "Concern really relates to scares around cyber security breaches" |
| Exploitation, stigmatisation and repercussions | Psychological trauma | "...it can add to the trauma of your diagnosis reading data without context or guidance." |

### Factors influencing willingness to share

Differences between support for sharing data and several respondent characteristics were explored (Appendix 1). We observed that respondents who reported they generally trusted two or more stakeholders were more likely to support sharing human research data with medical doctors (95% vs 82%, RD: 13%, 95% CI: 1-36%) and non-profit researchers (95% vs 75%, RD: 20%, 95% CI: 5-44%) than those who didn’t. Respondents who reported they generally trusted two or more stakeholders were also more likely to consent to researchers sharing their anonymised treatment data with medical doctors (99% vs 88%, RD: 11%, 95% CI: 2-34%) and to us sharing survey data publicly (81% vs 56%, RD: 25%, 95% CI: 4-49%). However, beyond this we did not observe any meaningful differences between attitudes of respondents according to their prior participation in research, successful completion of the comprehension check, familiarity with the topic of data sharing or the order in which the reasons for and against sharing were displayed.

## DISCUSSION

### Principal findings

In this study we explored what Australians affected by cancer think about oncology researchers sharing research data with third parties, including the public. This is in contrast with the subtly different, and more commonly investigated questions of how willing people are to donate their medical information to research, and what they think about researchers accessing routinely collected health data. Our study found that most respondents agreed that oncology researchers should share their research data with medical and non-profit researchers (90-95%) but noted less support for sharing with for-profit researchers (64%) and the public (61%). However, interestingly, attitudes did not appear to vary according to whether data are sourced from human participants or not. Very similar findings were also observed when respondents were asked about cancer researchers sharing de-identified research data containing their treatment information. Additionally, after being shown a visualisation of the data, 80% of respondents supported the public release of de-identified survey data.

The observation of high levels of support for medical researchers making data available to clinician and non-profit researchers, but not for sharing with the private sector, is consistent with other research [4,12,14,15,21]. For example, in the setting of clinical trials, a 2020 survey of 81 Australians affected by breast cancer showed that respondents were much more supportive of university and health agencies obtaining access to de-identified trial data than for-profit companies [4]. Similarly, a survey of 771 clinical trial participants in the United States, noted that 93% of respondents supported sharing their clinical trial data with academic and other not-for-profit researchers, whereas 82% supported sharing with companies developing medical products [15]. We also see similar discrepancies in the context of omics data as well. For example, Barnes and colleagues [14] observed support for sharing data collected to develop a proteomic test for stroke with non-profit and for-profit organisations decline from 87% to 53% respectively.

### Potential implications of our findings

Concerns among patients and the public regarding misuse, re-identification and loss of control over their information could be allayed by the following strategies. One strategy to balance the desire or need to make human research data publicly available and concerns about third parties profiting from research data, could be the application of non-commercial public copyright licenses (e.g., CC BY-NC 4.0), along with an explanation to participants that while data will be made public it cannot be legally used for commercial purposes. However, ambiguities in the definition of ‘non-commercial’, as well as the possible negative impacts of applying such licenses on data utility would also need to be considered with this approach [22]. A further strategy to address these concerns would be to submit data to FAIR-compliant, controlled-access repositories, where requests are reviewed to ensure they are in line with both the research community’s and participants’ values (e.g., methodologically sound, non-commercial motives).

Importantly, our study also suggests that visual aids could be a useful tool in low-risk research settings to help prospective participants better understand exactly what data will be collected and reused after the conclusion of the study. This strategy may also reduce anxieties about re-identification when researchers need to obtain consent to make data available to third parties. Both are findings that have been observed in other consent settings [23].

### Strengths and limitations of the study

Our study has several strengths which give us confidence that we have been able to elicit robust, informed, and high integrity opinions on the topic of data sharing. Firstly, our study was more than three times larger than a previous survey of Australian breast cancer patients’ attitudes on data sharing done in 2020 [4]. Our study also included several educational materials to help explain concepts like de-identification that are typically not well understood [21,24], and incorporated a comprehension check and several fraud detection mechanisms. However, there are some limitations. The use of public advertisements and convenience sampling means we are unable to determine the non-response rate, nor assess possible response biases. For example, our decision to obtain consent to make de-identified survey data available on request may have deterred people with strong views against sharing from participating. Further, the decision to host the survey online, and in English, limited our potential pool of participants. We also received five times more responses to the survey from women than men. However, prior research suggests that this is unlikely to have impacted the generalisability of our findings [15,25].

## CONCLUSION

Our survey has shown that Australians affected by cancer support the sharing of both human and non-human research data, particularly with clinician and non-profit researchers. Visualisations of the data to be shared may also help increase comprehension and decrease anxiety when obtaining consent to share de-identified research data publicly. Overall, these results should help alleviate any concerns about research participants’ attitudes on data sharing, as well as boost researchers’ motivation for sharing.

## Data Availability

Aggregate data, study materials and analytic code are publicly available on the project's Open Science Framework page under a Creative Commons Zero v1.0 Universal (CC0 1.0) license (DOI: 10.17605/OSF.IO/M52K3). Furthermore, so as not to deter people with strong views against public sharing of data from participating in the survey, we did not seek participants' consent to share de-identified participant-level data publicly. However, we did obtain participants' consent to release de-identified data to researchers for future research. Researchers interested in obtaining access to the study dataset can do so by following the instructions on the project's Open Science Framework page (https://osf.io/m52k3/).

https://doi.org/10.17605/OSF.IO/M52K3

## Disclosures

### Availability of data and materials

Aggregate data, study materials and analytic code have been posted on the project’s Open Science Framework page under a Creative Commons Zero v1.0 Universal (CC0 1.0) license (DOI: 10.17605/OSF.IO/M52K3). Furthermore, so as not to deter people with strong views against public sharing of data from participating in the survey, we did not seek participants’ consent to share de-identified participant-level data publicly. However, we did obtain participants’ consent to release de-identified data to researchers for future research. Researchers interested in obtaining access to the study dataset can do so by following the instructions on the project’s Open Science Framework page (https://osf.io/m52k3/).

## Acknowledgements

We thank Albertine Hamilton, Ellie Freeman, Claudia Hooper, Daniel Sapkaroski, Michael MacManus, Nigel Anderson, Farshad Foroudi, the Peter MacCallum Cancer Centre Wellbeing Centre and Head and Neck Cancer Australia for their assistance with recruitment. Some participants in this research were also recruited from Breast Cancer Network Australia’s (BCNA) Review and Survey Group, a national, online group of Australian women living with breast cancer who are interested in receiving invitations to participate in research. We acknowledge the contribution of the women involved in the Review and Survey Group who participated in this project. We also thank A/Prof Sue Finch for her advice on the statistical analysis.

## References

1. Foley C. How the United Nations’ new ‘open science framework’ could speed up the pace of discovery. The Conversation, 2021. December 4th, 2021. https://theconversation.com/how-the-united-nations-new-open-science-framework-could-speed-up-the-pace-of-discovery-173148. [Last accessed: February 27th, 2023.]

2. National Health and Medical Research Council (NHMRC). NHMRC’s revised Open Access Policy released. September 20th, 2022. https://www.nhmrc.gov.au/about-us/news-centre/nhmrcs-revised-open-access-policy-released. [Last accessed: March 8th, 2023.]

3. Research Australia. Public Opinion Poll on Health and Medical Research & Innovation. December 9th, 2020. https://issuu.com/researchaustralia/docs/2020_opinion_poll_report_final. [Last accessed: March 8th, 2023.]

4. Hutchings E, Butcher BE, Butow P, Boyle FM. Attitudes of Australian breast cancer patients toward the secondary use of administrative and clinical trial data. Asia Pac J Clin Oncol 2022. DOI: 10.1111/ajco.13734.

5. Errington TM, Denis A, Perfito N, Iorns E, Nosek BA. Challenges for assessing replicability in preclinical cancer biology. eLife. 2021; 10: e67995. DOI: 10.7554/eLife.67995.

6. Shahin MH, Bhattacharya S, Silva D, Kim S, Burton J, Podichetty J, et al. Open Data Revolution in Clinical Research: Opportunities and Challenges. Clinical and Translational Science. 2020; 13(4): 665–74. DOI: 10.1111/cts.12756.

7. Walters C, Harter ZJ, Wayant C, Vo N, Warren M, Chronister J, et al. Do oncology researchers adhere to reproducible and transparent principles? A cross-sectional survey of published oncology literature. BMJ Open 2019; 9(12): e033962. DOI: 10.1136/bmjopen-2019-033962.

8. Hamilton DG, Page MJ, Finch S, Everitt S, Fidler F. How often do cancer researchers make their data and code available and what factors are associated with sharing? BMC Medicine 2022 Nov 9; 20(1): 438. DOI: 10.1186/s12916-022-02644-2.

9. Zuiderwijk A, Shinde R, Jeng W. What drives and inhibits researchers to share and use open research data? A systematic literature review to analyze factors influencing open research data adoption. PLOS ONE. 2020; 15(9): e0239283. DOI: 10.1371/journal.pone.0239283.

10. Hutchings E, Loomes M, Butow P, et al. A systematic literature review of researchers’ and healthcare professionals’ attitudes towards the secondary use and sharing of health administrative and clinical trial data. Syst Rev 2020; 9: 240. DOI: 10.1186/s13643-020-01485-5.

11. Howe N, Giles E, Newbury-Birch D, McColl E. Systematic review of participants’ attitudes towards data sharing: a thematic synthesis. J Health Serv Res Policy 2018; 23(2): 123–33. DOI: 10.1177/1355819617751555.

12. Broes S, Verbaanderd C, Casteels M, Lacombe D, Huys I. Sharing of Clinical Trial Data and Samples: The Cancer Patient Perspective. Front. Med. 2020; 7: 33. DOI: 10.3389/fmed.2020.00033.

13. Shabani M, Bezuidenhout L, Borry P. Attitudes of research participants and the general public towards genomic data sharing: a systematic literature review. Expert Review of Molecular Diagnostics. 2014; 14(8): 1053–65. DOI: 10.1586/14737159.2014.961917.

14. Barnes R, Votova K, Rahimzadeh V, Osman N, Penn AM, Zawati MH, et al. Biobanking for Genomic and Personalized Health Research: Participant Perceptions and Preferences. Biopreservation and Biobanking. 2020; 18(3): 204–12. DOI: 10.1089/bio.2019.0090.

15. Mello MM, Lieou V, Goodman SN. Clinical Trial Participants’ Views of the Risks and Benefits of Data Sharing. The New England Journal of Medicine 2018; 378(23): 2202–11. DOI: 10.1056/NEJMsa1713258.

16. Hamilton DG, Hong K, Fraser H, Rowhani-Farid A, Fidler F, Page MJ. Rates and predictors of data and code sharing in the medical and health sciences: A systematic review with meta-analysis of individual participant data. medRxiv 2023. DOI: 10.1101/2023.03.22.23287607.

17. Hamilton, DG. What Do Australians Affected by Cancer Think about Sharing Research Data? Open Science Framework 2023; DOI 10.17605/OSF.IO/strk9.

18. Sharma A, Minh Duc NT, Luu Lam, et al. A Consensus-Based Checklist for Reporting of Survey Studies (CROSS). J Gen Intern Med. 2021; 36(10): 3179–87. DOI: 10.1007/s11606-021-06737-1.

19. Kawano N, Nagahiro Y, Yoshida S, et al. Clinical features and treatment outcomes of opportunistic infections among human T-lymphotrophic virus type 1 (HTLV-1) carriers and patients with adult T-cell leukemia-lymphoma (ATL) at a single institution from 2006 to 2016. Journal of Clinical and Experimental Hematopathology. 2019; 59(4): 156–67. DOI: 10.3960/jslrt.18032.

20. McLaughlin GH. SMOG Grading-a New Readability Formula. Journal of Reading. 1969; 12(8): 639–46. https://www.jstor.org/stable/40011226.

21. Middleton A, Milne R, Almarri MA, Anwer S, Atutornu J, Baranova EE, et al. Global Public Perceptions of Genomic Data Sharing: What Shapes the Willingness to Donate DNA and Health Data? The American Journal of Human Genetics. 2020; 107(4): 743–52. DOI: 10.1016/j.ajhg.2020.08.023.

22. Matthews T. For Open Data, think twice before applying Non-Commercial conditions. Springer Nature Research Data Community. July 18th, 2022. https://go.nature.com/3cnFV5r.

23. Stewart JA, Wood L, Wiener J, Kennedy GD, Chu DI, Lancaster JR, et al. Visual teaching aids improve patient understanding and reduce anxiety prior to a colectomy. The American Journal of Surgery. 2021; 222(4): 780–5. DOI: 10.1016/j.amjsurg.2021.01.029.

24. Corman A, Canaway R, Culnane C, Teague V. Public comprehension of privacy protections applied to health data shared for research: An Australian cross-sectional study. International Journal of Medical Informatics 2022; 167: 104859. DOI: 10.1016/j.ijmedinf.2022.104859.

25. King T, Brankovic L, Gillard P. Perspectives of Australian adults about protecting the privacy of their health information in statistical databases. International Journal of Medical Informatics. 2012; 81(4): 279–89. DOI: 10.1016/j.ijmedinf.2012.01.005.

