## Appendix 1 for "What do Australians affected by cancer think about oncology researchers sharing research data: a cross-sectional survey"

### APPENDIX 1. ADDITIONAL STUDY INFORMATION.

#### Supplementary Methods

A short, cross-sectional survey designed to capture the views on data sharing of Australians affected by cancer was advertised on the University of Melbourne's website (https://science.unimelb.edu.au/about/news/your-data-your-say) on October 27th, 2022. Any person over the age of 18, who was an Australian citizen or resident, able to comprehend English, and had been previously diagnosed with a cancer of any kind was eligible to participate. Further information on the study methods is included on the project's Open Science Framework page. The findings of this study are reported in accordance with the Checklist for Reporting of Survey Studies (CROSS) guidelines (Supplementary Table 3). The study protocol for this study was not publicly registered prior to commencement.

##### Survey design

The survey contained 23 questions separated into five sections. The first section collected relevant demographic information from participants including: referral location, age, gender, rurality, level of education, diagnosed cancer, time since last treatment, prior research participation and trust in Australian healthcare stakeholders (i.e., their GP, other medical doctors, academic researchers, for-profit researchers and the Australian Government). The second explained the concept of 'research data' and the commonly stated reasons for and against sharing, with the order of reasons being randomised for each participant, to allow us to verify that order of reasons did not impact participant's attitudes towards data sharing. The third tested participants' comprehension of our definition of 'research data' using real-world data from a publication investigating viral and fungal infections in lymphoma patients [1], as well as captured whether participants were familiar with these concepts. The fourth characterised participants' general views on both the sharing of human and non-human research data, and the hypothetical sharing of their medical information with four different groups (medical doctors, non-profit researchers, for-profit researchers and the public). The fifth and final section provided a visual representation of what the survey data would look like, incorporating each participant's responses into one of the rows (see Figure 1), then proceeded to ask the participant for their views on: i) whether and how long the data should be retained, ii) whether they would hypothetically allow the research team to share it publicly, and iii) if not, which data, if any, they wished to be withheld and the reasons why.

Where possible, we used previously validated instruments relevant to an Australian setting to measure key demographics (e.g., rurality was measured using the Rural, Remote and Metropolitan Areas Classification [2], level of education was measured using the Australian Bureau of Statistics' (ABS) Australian Standard Classification of Education (ASCED) [3]) (refer to Supplementary Table 1 for further information). Furthermore, to avoid extensive piloting phases, as well as to allow comparison with relevant research, we adapted definitions, terminologies, and question structures from previous studies in the area [4,5]. For example, questions concerning trust in institutions, the recipients of the data, as well as familiarity with the topic were adapted from Middleton and colleagues who, along with other research groups, demonstrated these factors influence Australians' attitudes towards sharing in the setting of omics data [5,6,7]. More detailed information on the survey design, including the complete transcript, can be found on the project's Open Science Framework (OSF) page [Error! Reference source not found.].

Survey piloting and distribution

The survey was pilot tested between August and September 2022 by ten individuals of varying demographics (Supplementary Table 2). Piloters were asked to provide feedback on the survey, particularly the accessibility of the language, the onerousness of the questions, and the time taken to complete the survey. Further information on the piloting phase, including the feedback and subsequent revisions can be found on the project's OSF page. Following this, the readability of the survey transcript was then assessed using the SMOG (Simple Measure of Gobbledygook) scale [9] and further language edits were made to ensure that average readability score across the survey stayed at seventh grade reading level.

Qualtrics Solutions' Online Survey Software (Qualtrics, Provo, UT) was used to create and host the survey and three passive recruitment strategies were used to advertise the survey between October 27$^{th}$, 2022 and February 27$^{th}$, 2023. Specifically, the study was promoted: i) on various social media platforms, ii) within internal newsletters of cancer organisations, patient advocacy and support groups and patient accommodation services and iii) on physical flyers posted in several local cancer clinics. Several features were also incorporated into the survey to minimise data entry errors and prevent or identify submission of fraudulent responses. To detect and prevent fraudulent responses, first no financial incentives were offered to participate in the survey and the survey URL was not directly included in any of the advertisements. Second, Google reCAPTCHA codes, RelevantID® fraud detection checks and use of browser cookies to detect duplicate submissions were added. Survey completion times that were two standard deviations faster than the average piloter were also flagged. Finally, a series of manual data quality checks were also performed, which included screening for: duplicate answers to the long-form questions and discrepancies between survey start dates, reported referral avenues and advertisement initiation dates. Any respondent who failed any of these checks were categorised as suspected fraud.

Statistical analysis

Given the exploratory nature of the project, we set a target sample size of at least 100 responses to the survey. This size is consistent with, and larger than previous online surveys of cancer patients using similar survey tools and recruitment strategies [10,11,12,13]. Continuous data are presented as medians and interquartile ranges (IQR) and categorical data are presented as counts and proportions. Risk differences (RD) between proportions and associated 95% confidence intervals (CI) were calculated to assess differences in support for data sharing according to i) human versus non-human data type, ii) trust in multiple stakeholders, iii) previous participation in research, iv) comprehension of, and v) familiarity with data sharing, and vi) the order in which respondents were shown the reasons for and against sharing. Confidence intervals for rate differences between paired and independent binomial proportions were calculated using the method proposed by Tango and Newcombe respectively [14,15].

For responses to the open-ended questions, deductive content analysis was used to analyse responses. Specifically, a deductive approach to coding was used where responses were independently read and coded by two authors in accordance with the themes identified by the review of research participants' attitudes to data sharing by Howe and colleagues [16], with discrepancies resolved via discussion. The study codebook and inter-coder reliability statistics (unweighted kappa coefficients and percentage agreement) are available on the project's OSF page. Once all responses had been coded, we then calculated the frequency with which themes occurred in responses.

Quantitative analyses were performed in R (R Foundation for Statistical Computing, Vienna, Austria, v4.2.1) and qualitative analyses were performed in NVivo (QSR International Pty Ltd., v12.4.0). Bar charts presented in the main manuscript were created in Keynote and bar charts presented in the supplementary material were created in R using the HH package (v3.1-49) [17]. We did not attempt to impute missing data, nor did we use any methods to adjust for the potential non-representativeness of the sample. Missing responses are omitted from all reported results. Results concerning attitudes to sharing are also limited to respondents who correctly answered the comprehension check unless specified otherwise.

Ethics approval

The study protocol was reviewed and approved by the University of Melbourne's STEMM2 Human Ethics Committee (Ethics ID: 2022-22111-32090-5) prior to survey piloting and recruitment.

References

1. Kawano N, Nagahiro Y, Yoshida S, Tahara Y, Himeji D, Kuriyama T, et al. Clinical features and treatment outcomes of opportunistic infections among human T-lymphotrophic virus type 1 (HTLV-1) carriers and patients with adult T-cell leukemia-lymphoma (ATL) at a single institution from 2006 to 2016. Journal of Clinical and Experimental Hematopathology. 2019;59(4):156-67.
2. Department of Primary Industries and Energy, Department of Human Services and Health Rural, Remote and Metropolitan Areas Classification 1991 Census Edition. [(accessed on 18 October 2021)];1994 Available online: https://www.pc.gov.au/inquiries/completed/nursing-home-subsidies/submissions/subdr096/subdr096.pdf.
3. Australian Bureau of Statistics. *Australian Standard Classification of Education (ASCED)* [Internet]. Canberra: ABS; 2001 [cited 2023 April 3]. Available from: https://www.abs.gov.au/statistics/classifications/australian-standard-classification-education-asced/latest-release.
4. Mello MM, Lieou V, Goodman SN. Clinical Trial Participants' Views of the Risks and Benefits of Data Sharing. The New England Journal of Medicine; 2018 Jun 7;378(23):2202-11.
5. Middleton A, Milne R, Almarri MA, Anwer S, Atutornu J, Baranova EE, et al. Global Public Perceptions of Genomic Data Sharing: What Shapes the Willingness to Donate

DNA and Health Data? The American Journal of Human Genetics. 2020 Oct 1;107(4):743-52.
6. Lynch F, Meng Y, Best S, Goranitis I, Savulescu J, Gyngell C, et al. Australian public perspectives on genomic data storage and sharing: Benefits, concerns and access preferences. Eur J Med Genet. 2023 Jan;66(1):104676.
7. Braunack-Mayer A, Fabrianesi B, Street J, O'Shaughnessy P, Carter SM, Engelen L, et al. Sharing Government Health Data With the Private Sector: Community Attitudes Survey. J Med Internet Res. 2021 Oct 1;23(10):e24200.
8. Hamilton, DG. What Do Australians Affected by Cancer Think about Sharing Research Data? Open Science Framework 2023; DOI 10.17605/OSF.IO/strk9.
9. McLaughlin GH. SMOG Grading-a New Readability Formula. Journal of Reading. 1969;12(8):639-46. https://www.jstor.org/stable/40011226
10. Lapedis CJ, Horowitz JK, Brown L, Tolle BE, Smith LB, Owens SR. The Patient-Pathologist Consultation Program: A Mixed-Methods Study of Interest and Motivations in Cancer Patients. Archives of Pathology & Laboratory Medicine 2019;144:490-6. https://doi.org/10.5858/arpa.2019-0105-OA.
11. Hashmi F, Gregor N, Liszewski B, Bola R, Kulczyski S, Nathoo D, et al. It Only Takes a Minute: The Development and Implementation of a Patient Experience Survey in Radiation Therapy. Journal of Medical Imaging and Radiation Sciences 2019;50:5-11. https://doi.org/10.1016/j.jmir.2018.07.006.
12. Woudstra AJ, Smets EMA, Dekker E, Broens THF, Penning J, Smith S, et al. Development and pilot-testing of a colorectal cancer screening decision aid for individuals with varying health literacy levels. Patient Education and Counseling 2019;102:1847-58. https://doi.org/10.1016/j.pec.2019.04.029.
13. Markovic C, Mackenzie L, Lewis J, Singh M. Working with cancer: A pilot study of work participation among cancer survivors in Western Sydney. Aust Occup Ther J 2020;67:592-604. https://doi.org/10.1111/1440-1630.12685.
14. Tango T. Equivalence test and confidence interval for the difference in proportions for the paired-sample design. Statistics in Medicine 1998; 17:891-908.
15. Newcombe RG. Interval estimation for the difference between independent proportions: comparison of eleven methods. Statistics in Medicine 1998.
16. Howe N, Giles E, Newbury-Birch D, McColl E. Systematic review of participants' attitudes towards data sharing: a thematic synthesis. J Health Serv Res Policy. 2018 Apr;23(2):123-33.
17. Heiberger RM, Robbins NB. Design of Diverging Stacked Bar Charts for Likert Scales and Other Applications. Journal of Statistical Software 2014; 57(5): 1-32. https://www.jstatsoft.org/v57/i05/.
18. Hutchings E, Butcher BE, Butow P, Boyle FM. Attitudes of Australian breast cancer patients toward the secondary use of administrative and clinical trial data. Asia Pac J Clin Oncol 2022. DOI: 10.1111/ajco.13734.

**Supplementary Table 1. Instruments used in the design of the survey.**

| Section | Question(s) | Theme(s) | Instrument(s) used |
|---|---|---|---|
| Demographics | Q1-Q4 | Referral source; Age; Gender; Australian citizenship/residency | Question wordings were designed by the authors. |
| | Q5 | Rurality | Rural, Remote and Metropolitan Areas Classification 1994 [2]. |
| | Q6 | Education history | Australian Bureau of Statistics' (ABS) Australian Standard Classification of Education (ASCED) [3] |
| | Q7 | Diagnosed cancer | Categories were adapted from the Cancer Council Victoria (https://www.cancervic.org.au/cancer-information/types-of-cancer) |
| | Q8-Q9 | Time since treatment; Research participation | Question wordings were designed by the authors. |
| | Q10 | Trust | Question wording was adapted from Middleton et al [5]. |
| Background | - | Definition of data | The wording and visualisation of research data was designed by the authors. |
| | - | Reasons for and against sharing | The reasons were adapted from Mello et al [4]. |
| Knowledge | Q11 | Comprehension check | The comprehension check was created by the authors using research data published by Kawano et al [1]. |
| | Q12 | Familiarity with data sharing | Question wording was adapted from Middleton et al [5]. |
| | Q13 | Reasons for familiarity | Question wording was adapted from Middleton et al [5]. |
| General views | Q14 | Support for sharing human research data | Question wording was adapted from Middleton et al [5]. |
| | Q15 | Support for sharing non-human data | Question wording was adapted from Middleton et al [5]. |
| | Q16 | Estimation of data sharing | Question wording was adapted from Hamilton et al 2023 [19]. |
| | Q17 | Sharing of respondents' medical data | Question wording was adapted from Middleton et al [5]. |

| | | | |
|---|---|---|---|
| Survey data | Q18-21 | Survey data deletion;<br>Survey data retention;<br>Survey data sharing; | All question wordings and visualisation of research data were designed by the authors. |
| | Q22-23 | Concerns about sharing;<br>Other thoughts | All question wordings were designed by the authors. Codebook developed by Howe et al [16] were used to perform qualitative analyses |

**Supplementary Table 2. Characteristics of all respondents.**

| | Piloters | | Survey | | Hutchings 2022 | |
|---|---|---|---|---|---|---|
| | N | % | N | % | N | % |
| Sex | | | | | | |
| - Male | 4 | 40% | 87 | 16% | 1 | 1% |
| - Female | 6 | 60% | 457 | 84% | 131 | 99% |
| Age | | | | | | |
| - Median (IQR) | 54 | 44 to 72 | 66 | 59 to 73 | NA | NA |
| Rurality | | | | | | |
| - Metropolitan area | 6 | 60% | 330 | 62% | - | - |
| - Small or large rural town | 4 | 40% | 45 | 30% | - | - |
| - Remote or very remote community | 0 | 0% | 158 | 8% | - | - |
| Level of education | | | | | | |
| - Postgraduate degree | 3 | 30% | 140 | 26% | 44 | 33% |
| - Bachelor's degree | 1 | 10% | 122 | 23% | 41 | 31% |
| - Certificate or Diploma | 4 | 40% | 139 | 26% | | |
| - High school | 2 | 20% | 129 | 24% | 47 | 36% |
| - Primary school | 0 | 0% | 5 | 1% | | |
| Diagnosed cancer | | | | | | |
| - Breast cancer | 4 | 40% | 292 | 54% | 132 | 100% |
| - Melanoma of the skin | 1 | 10% | 30 | 6% | 0 | 0% |
| - Prostate cancer | 3 | 30% | 29 | 5% | 0 | 0% |
| - Colorectal cancer | 1 | 10% | 28 | 5% | 0 | 0% |
| - Other | 1 | 10% | 23 | 4% | 0 | 0% |
| Time since treatment | | | | | | |
| - Currently receiving treatment | 1 | 10% | 115 | 21% | 16 | 12% |
| - <1 year ago | 3 | 30% | 86 | 16% | 6 | 5% |
| - 1-5 years ago | 3 | 30% | 145 | 27% | 27 | 23% |
| - 6-10 years ago | 1 | 10% | 83 | 15% | 83 | 72% |
| - 10+ years ago | 1 | 10% | 109 | 20% | | |
| Participated in cancer research | | | | | | |
| - Yes (Clinical trial) | 2 | 20% | 49 | 9% | 31 | 24% |
| - Yes (Other research) | 0 | 0% | 156 | 29% | - | - |
| - No | 8 | 80% | 324 | 60% | - | - |
| - Unsure | 0 | 0% | 32 | 6% | - | - |

Note: 'Piloters' refers to the 10 individuals who tested the survey prior to activation, 'Survey' refers to all participants that contributed data to survey, and 'Hutchings et al 2022' refers to the characteristics of the sample of the study by Hutchings et al 2022 [18].

**Supplementary Table 3. Checklist for Reporting Of Survey Studies (CROSS). (Note: References are to sections of the Supplementary Methods)**

| Section/topic | Item | Item description | Reported on page # |
|---|---|---|---|
| **Title and abstract** | | | |
| Title and abstract | 1a | State the word "survey" along with a commonly used term in title or abstract to introduce the study's design. | Page 1 |
| | 1b | Provide an informative summary in the abstract, covering background, objectives, methods, findings/results, interpretation/discussion, and conclusions. | Page 1 |
| **Introduction** | | | |
| Background | 2 | Provide a background about the rationale of study, what has been previously done, and why this survey is needed. | Page 1 & 2 |
| Purpose/aim | 3 | Identify specific purposes, aims, goals, or objectives of the study. | Page 2 |
| **Methods** | | | |
| Study design | 4 | Specify the study design in the methods section with a commonly used term (e.g., cross-sectional or longitudinal). | Page 2 |
| Data collection methods | 5a | Describe the questionnaire (e.g., number of sections, number of questions, number and names of instruments used). | Page 2-3 |
| | 5b | Describe all questionnaire instruments that were used in the survey to measure particular concepts. Report target population, reported validity and reliability information, scoring/classification procedure, and reference links (if any). | Appendix 1 Table S1 |
| | 5c | Provide information on pretesting of the questionnaire, if performed (in the article or in an online supplement). Report the method of pretesting, number of times questionnaire was pre-tested, number and demographics of participants used for pretesting, and the level of similarity of demographics between pre-testing participants and sample population. | Page 2-3 |
| | 5d | Questionnaire if possible, should be fully provided (in the article, or as appendices or as an online supplement). | Appendix 2 |
| Sample characteristics | 6a | Describe the study population (i.e., background, locations, eligibility criteria for participant inclusion in survey, exclusion criteria). | Page 2 |
| | 6b | Describe the sampling techniques used (e.g., single stage or multistage sampling, simple random sampling, stratified sampling, cluster sampling, convenience sampling). Specify the locations of sample participants whenever clustered sampling was applied. | Appendix 1 |
| | 6c | Provide information on sample size, along with details of sample size calculation. | Appendix 1 |
| | 6d | Describe how representative the sample is of the study population (or target population if possible), particularly for population-based surveys. | Page 8 & Appendix 1 Table S2 |
| Survey administration | 7a | Provide information on modes of questionnaire administration, including the type and number of contacts, the location where the survey was conducted (e.g., outpatient room or by use of online tools, such as SurveyMonkey). | Page 2-3 |
| | 7b | Provide information of survey's time frame, such as periods of recruitment, exposure, and follow-up days. | Appendix 1 |

| | | | |
|---|---|---|---|
| | 7c | Provide information on the entry process:<br>->For non-web-based surveys, provide approaches to minimize human error in data entry.<br>->For web-based surveys, provide approaches to prevent "multiple participation" of participants. | Appendix 1 |
| Study preparation | 8 | Describe any preparation process before conducting the survey (e.g., interviewers' training process, advertising the survey). | Appendix 1 |
| Ethical considerations | 9a | Provide information on ethical approval for the survey if obtained, including informed consent, institutional review board [IRB] approval, Helsinki declaration, and good clinical practice [GCP] declaration (as appropriate). | Page 3 |
| | 9b | Provide information about survey anonymity and confidentiality and describe what mechanisms were used to protect unauthorized access. | Appendix 1 |
| Statistical analysis | 10a | Describe statistical methods and analytical approach. Report the statistical software that was used for data analysis. | Appendix 1 |
| | 10b | Report any modification of variables used in the analysis, along with reference (if available). | Not applicable |
| | 10c | Report details about how missing data was handled. Include rate of missing items, missing data mechanism (i.e., missing completely at random [MCAR], missing at random [MAR] or missing not at random [MNAR]) and methods used to deal with missing data (e.g., multiple imputation). | Appendix 1 |
| | 10d | State how non-response error was addressed. | Page 8 |
| | 10e | For longitudinal surveys, state how loss to follow-up was addressed. | Not applicable |
| | 10f | Indicate whether any methods such as weighting of items or propensity scores have been used to adjust for non-representativeness of the sample. | Appendix 1 |
| | 10g | Describe any sensitivity analysis conducted. | Page 3 |
| **Results** | | | |
| Respondent characteristics | 11a | Report numbers of individuals at each stage of the study. Consider using a flow diagram, if possible. | Figure S1 |
| | 11b | Provide reasons for non-participation at each stage, if possible. | Figure S1 |
| | 11c | Report response rate, present the definition of response rate or the formula used to calculate response rate. | Page 3 |
| | 11d | Provide information to define how unique visitors are determined. Report number of unique visitors along with relevant proportions (e.g., view proportion, participation proportion, completion proportion). | Appendix 1 & Figure S1 |
| Descriptive results | 12 | Provide characteristics of study participants, as well as information on potential confounders and assessed outcomes. | Page 4 |
| Main findings | 13a | Give unadjusted estimates and, if applicable, confounder-adjusted estimates along with 95% confidence intervals and p-values. | Page 4-5 |
| | 13b | For multivariable analysis, provide information on the model building process, model fit statistics, and model assumptions (as appropriate). | Not applicable |
| | 13c | Provide details about any sensitivity analysis performed. If there are considerable amount of missing data, report sensitivity analyses comparing the results of complete cases with that of the imputed dataset | Not applicable |

| | | | |
|---|---|---|---|
| | | (if possible). | |
| **Discussion** | | | |
| Limitations | 14 | Discuss the limitations of the study, considering sources of potential biases and imprecisions, such as non-representativeness of sample, study design, important uncontrolled confounders. | Page 8 |
| Interpretations | 15 | Give a cautious overall interpretation of results, based on potential biases and imprecisions and suggest areas for future research. | Page 7-8 |
| Generalizability | 16 | Discuss the external validity of the results. | Page 8 |
| **Other sections** | | | |
| Role of funding source | 17 | State whether any funding organization has had any roles in the survey's design, implementation, and analysis. | Page 8 |
| Conflict of interest | 18 | Declare any potential conflict of interest. | Page 8 |
| Acknowledgements | 19 | Provide names of organizations/persons that are acknowledged along with their contribution to the research. | Page 8 |

**Supplementary Figure 1. Proportion of missing data by survey question. (Note: questions not shown to all respondents are not included.)**

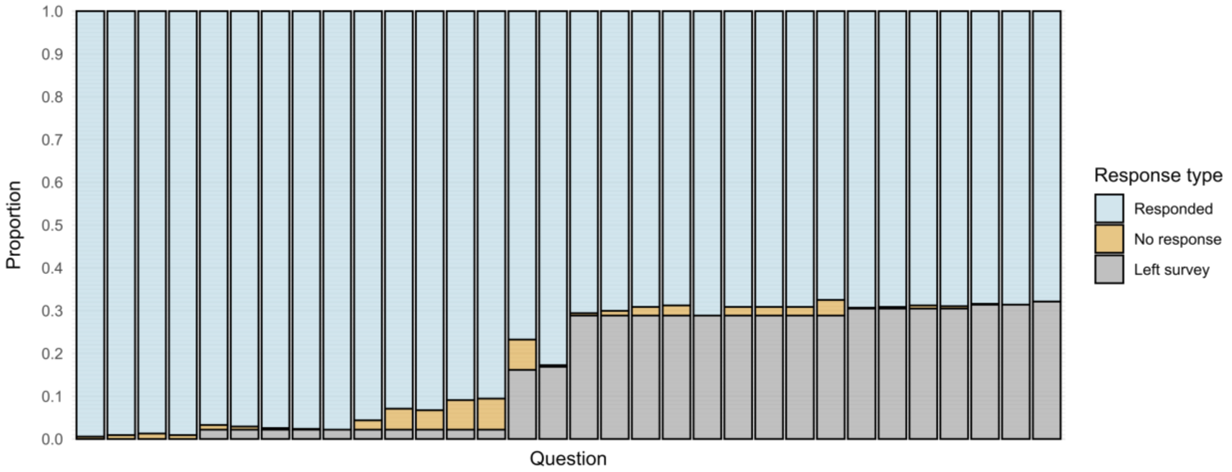


| Q1 | Q2 | Q3 | Q4 | Q5 | Q6 | Q7 | Q8 | Q9 | Q10-1 | Q10-2 | Q10-3 | Q10-4 | Q10-5 | Q11 | Q12 |
|---|---|---|---|---|---|---|---|---|---|---|---|---|---|---|---|
| 548 | 546 | 544 | 546 | 533 | 535 | 537 | 538 | 539 | 527 | 512 | 514 | 501 | 499 | 423 | 456 |
| 3 | 5 | 7 | 5 | 6 | 4 | 2 | 1 | 0 | 12 | 27 | 25 | 38 | 40 | 39 | 2 |
| 0 | 0 | 0 | 0 | 12 | 12 | 12 | 12 | 12 | 12 | 12 | 12 | 12 | 12 | 89 | 93 |

| Q14-1 | Q14-2 | Q14-3 | Q14-4 | Q15-1 | Q15-2 | Q15-3 | Q15-4 | Q16-1 | Q17-1 | Q17-2 | Q17-3 | Q17-4 | Q18 | Q19 | Q20 |
|---|---|---|---|---|---|---|---|---|---|---|---|---|---|---|---|
| 389 | 386 | 381 | 379 | 392 | 381 | 381 | 381 | 372 | 382 | 381 | 379 | 380 | 377 | 378 | 374 |
| 3 | 6 | 11 | 13 | 0 | 11 | 11 | 11 | 20 | 1 | 2 | 4 | 3 | 1 | 0 | 0 |
| 159 | 159 | 159 | 159 | 159 | 159 | 159 | 159 | 159 | 168 | 168 | 168 | 168 | 173 | 173 | 177 |

**Supplementary Figure 2. Respondents' views on the general sharing of research data derived from non-human and human participants by levels of trust in multiple stakeholders.**

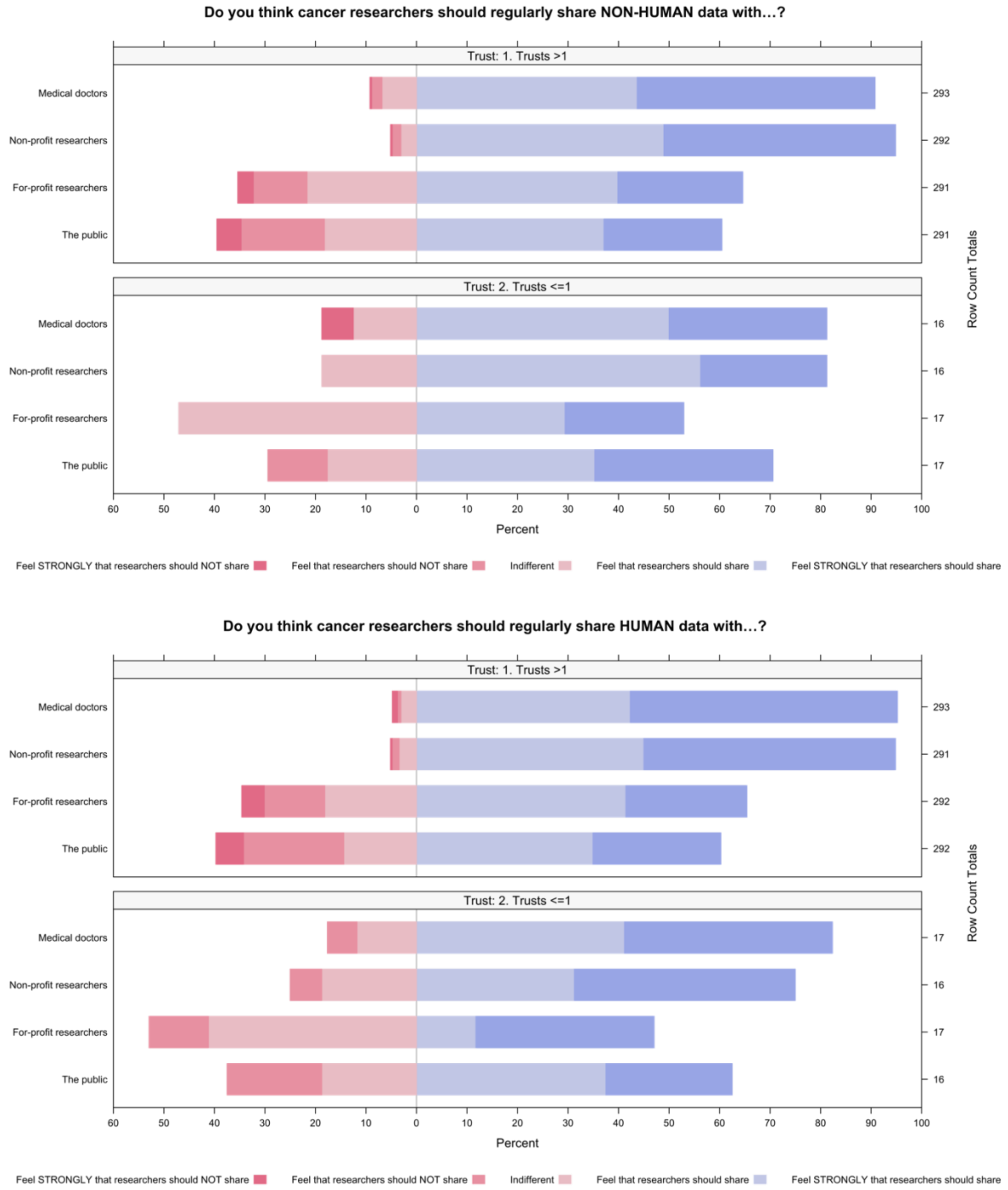

**Supplementary Figure 3. Respondents' views on the general sharing of research data derived from non-human and human participants by previous participation in research.**

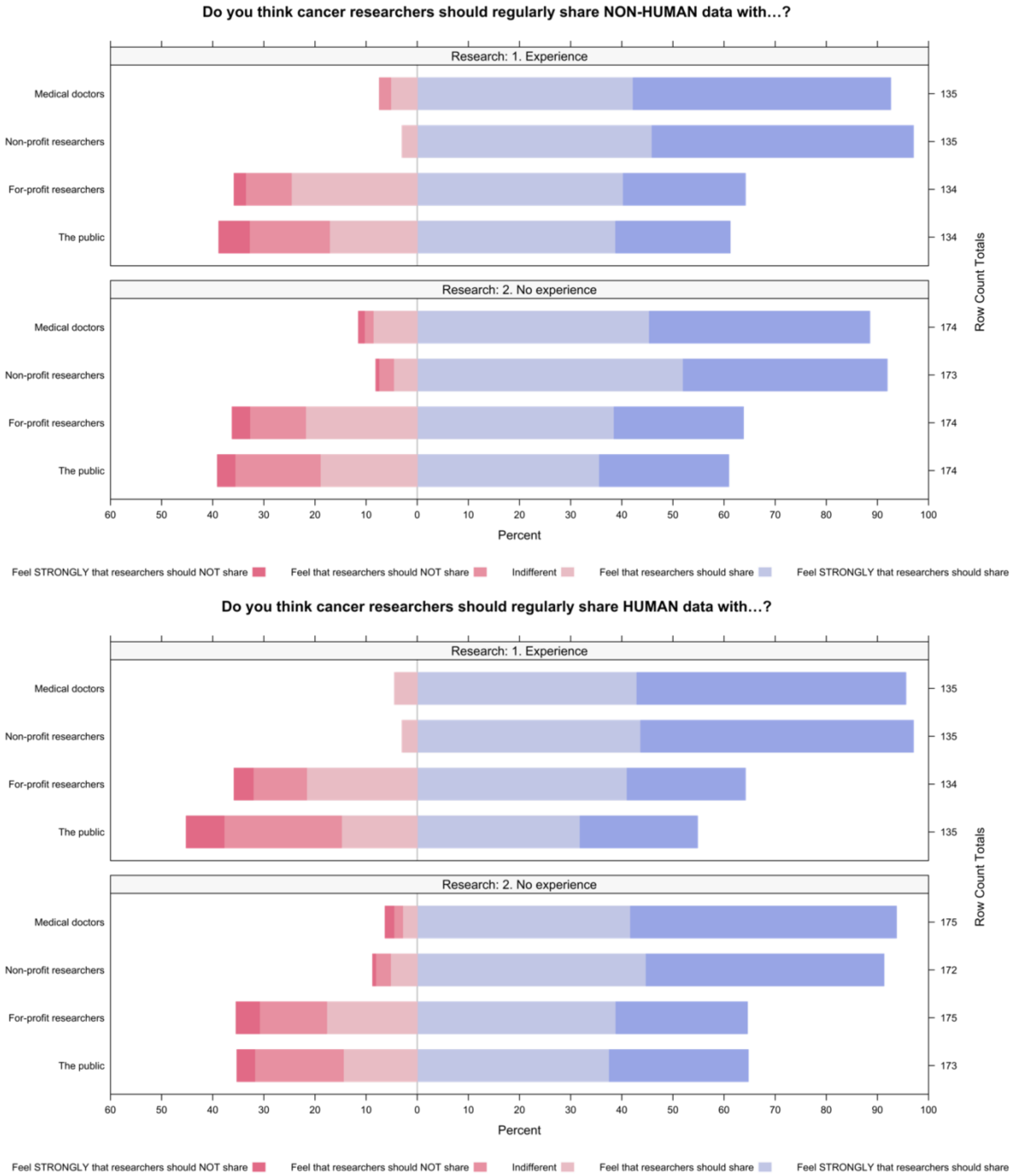

**Supplementary Figure 4. Respondents' views on the general sharing of research data derived from non-human and human participants by familiarity with data sharing.**

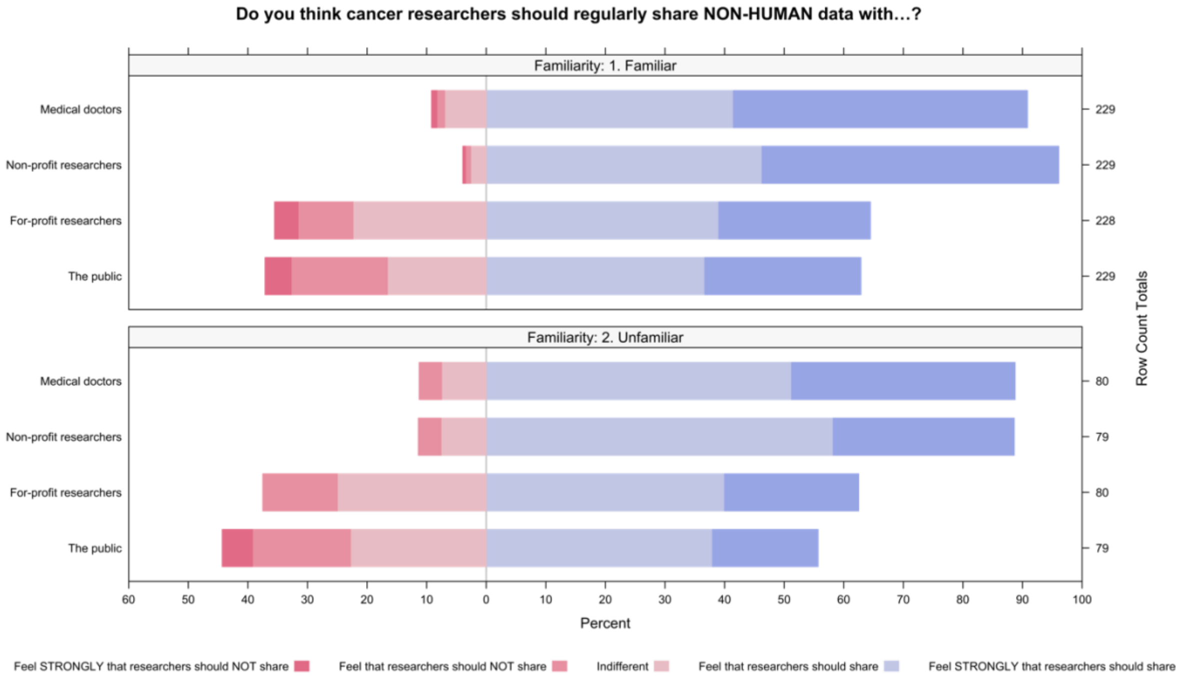


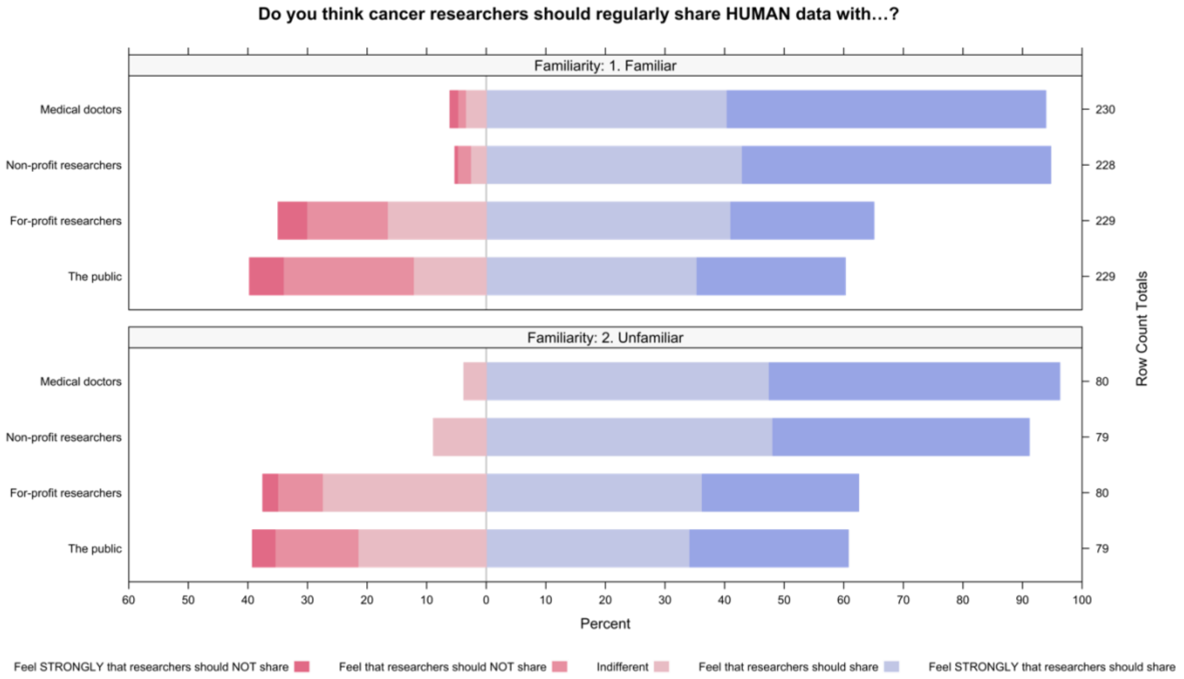

**Supplementary Figure 5. Respondents' views on the general sharing of research data derived from non-human and human participants by the order which participants were shown the advantages and disadvantages of sharing data.**

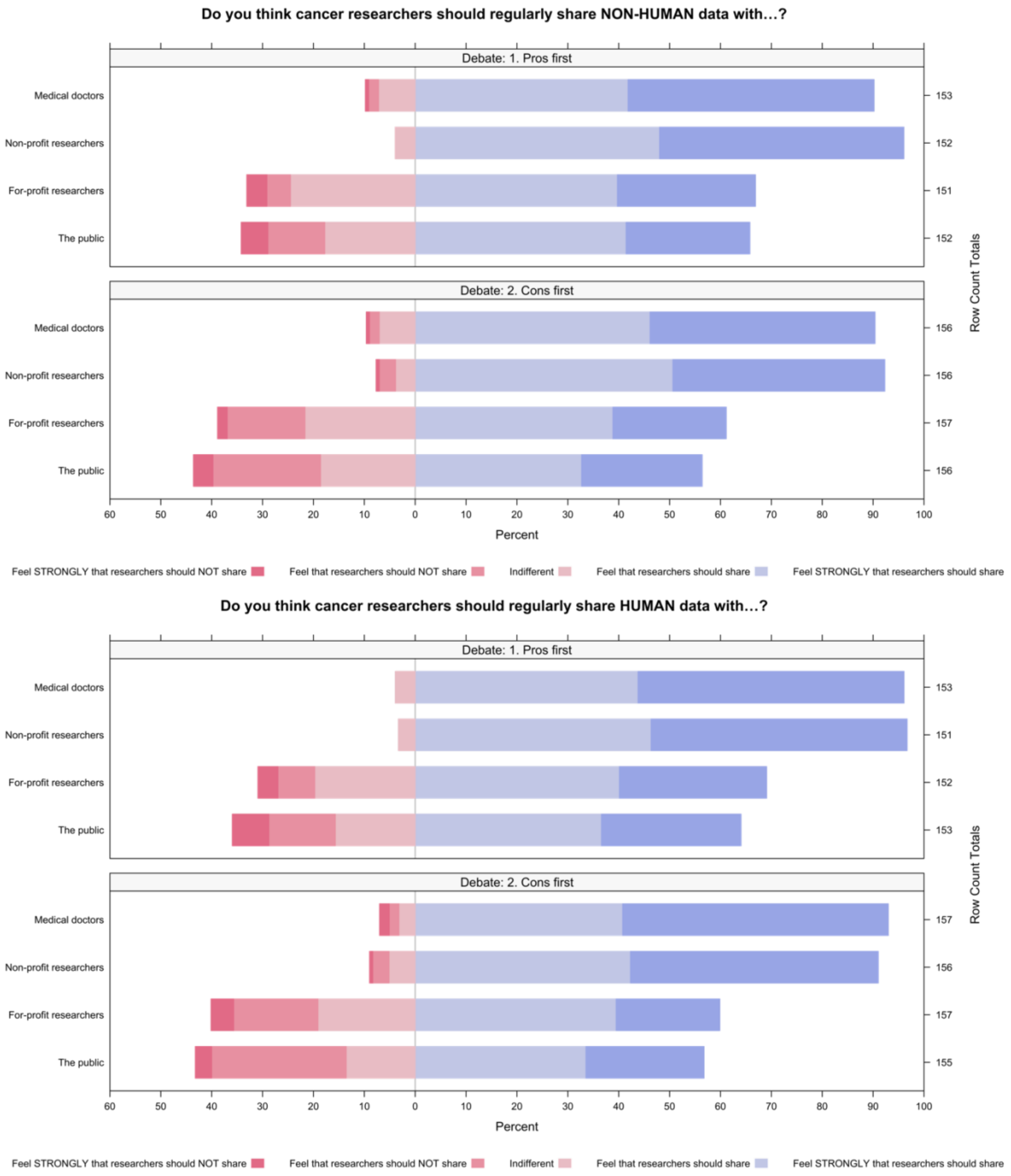

**Supplementary Figure 6. Respondents' views on the general sharing of research data derived from non-human and human participants by whether participants passed the comprehension check or not.**

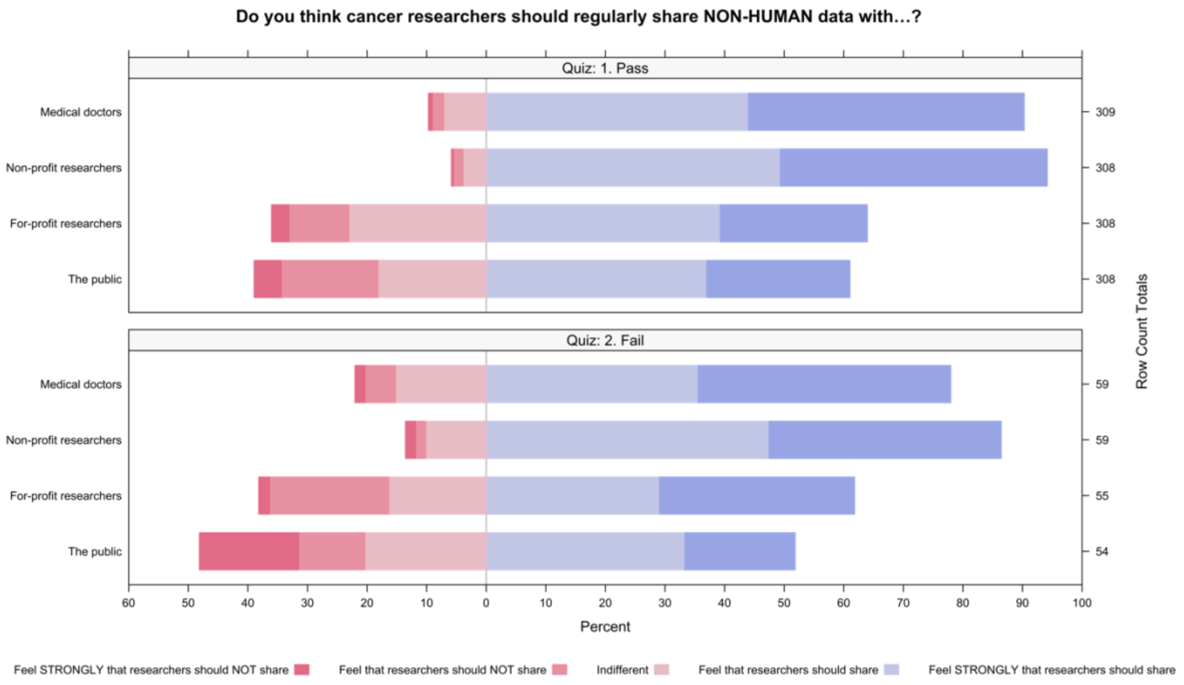


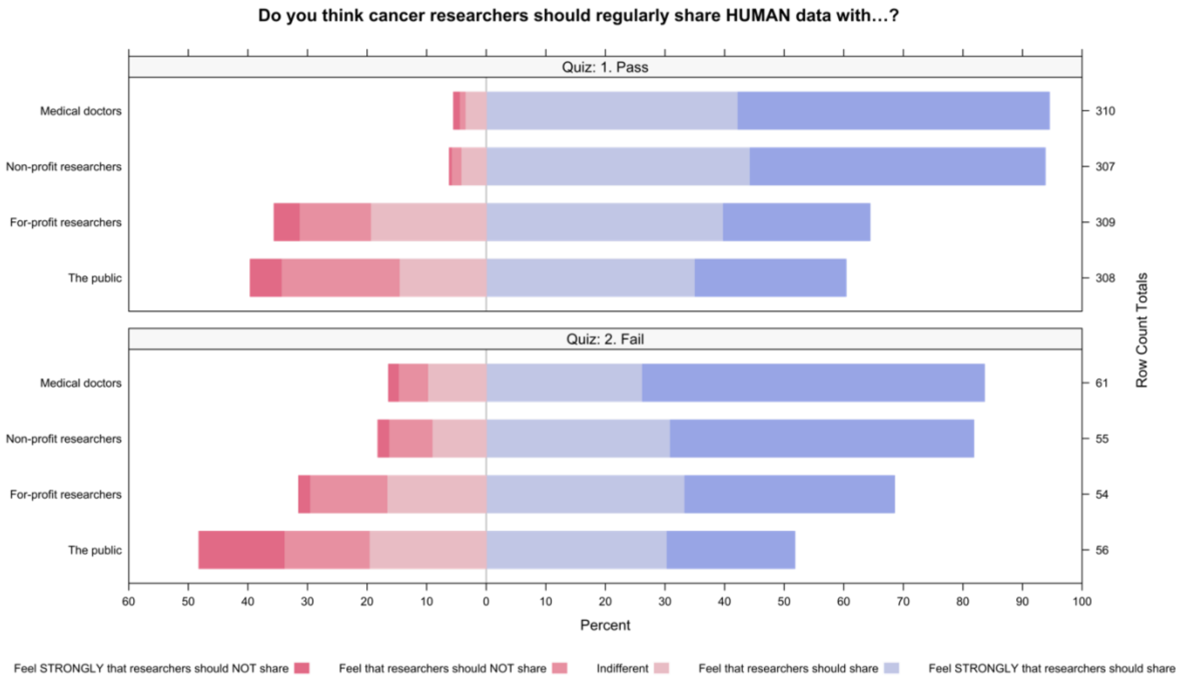

**Supplementary Figure 7. Respondents' views on researchers hypothetically sharing their treatment information by levels of trust in multiple stakeholders.**

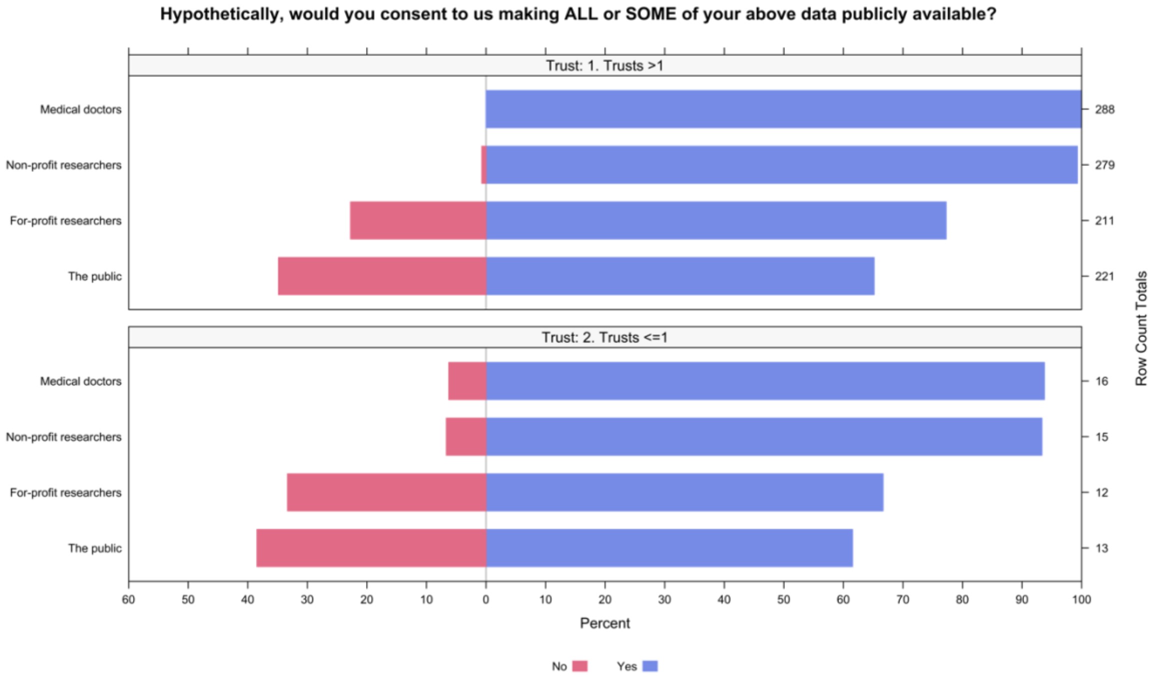


**Supplementary Figure 8. Respondents' views on researchers hypothetically sharing their treatment information by previous participation in research.**

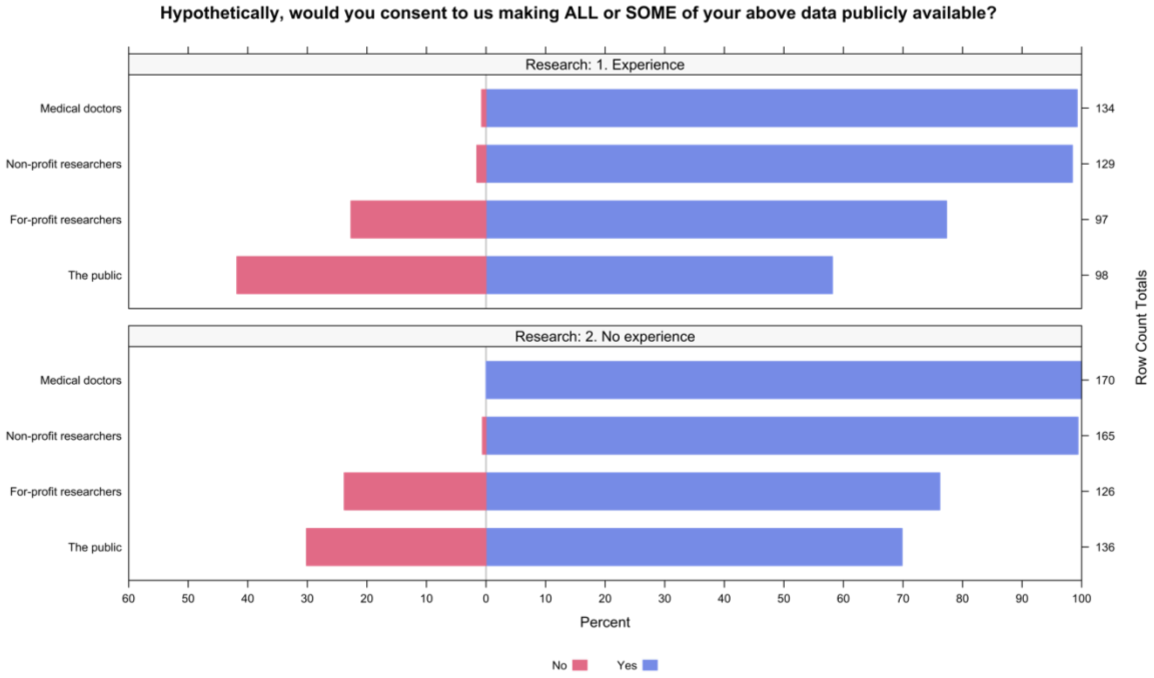

**Supplementary Figure 9. Respondents' views on researchers hypothetically sharing their treatment information by familiarity with data sharing.**

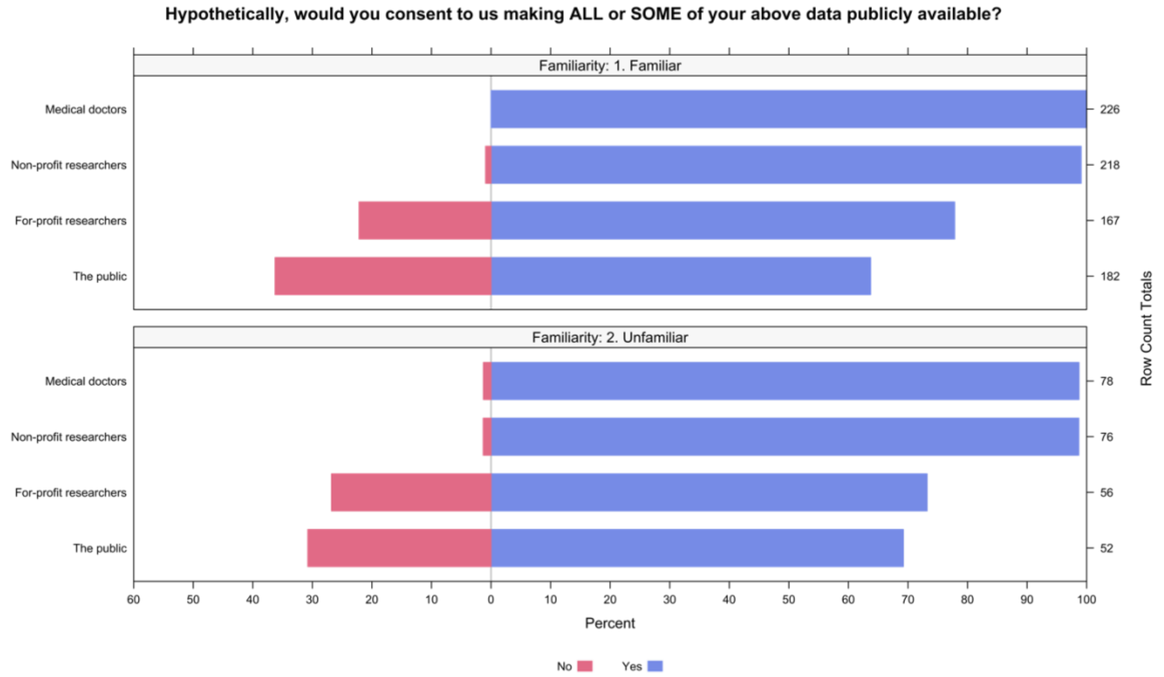


**Supplementary Figure 10. Respondents' views on researchers hypothetically sharing their treatment information by the order which participants were shown the advantages and disadvantages of sharing data.**

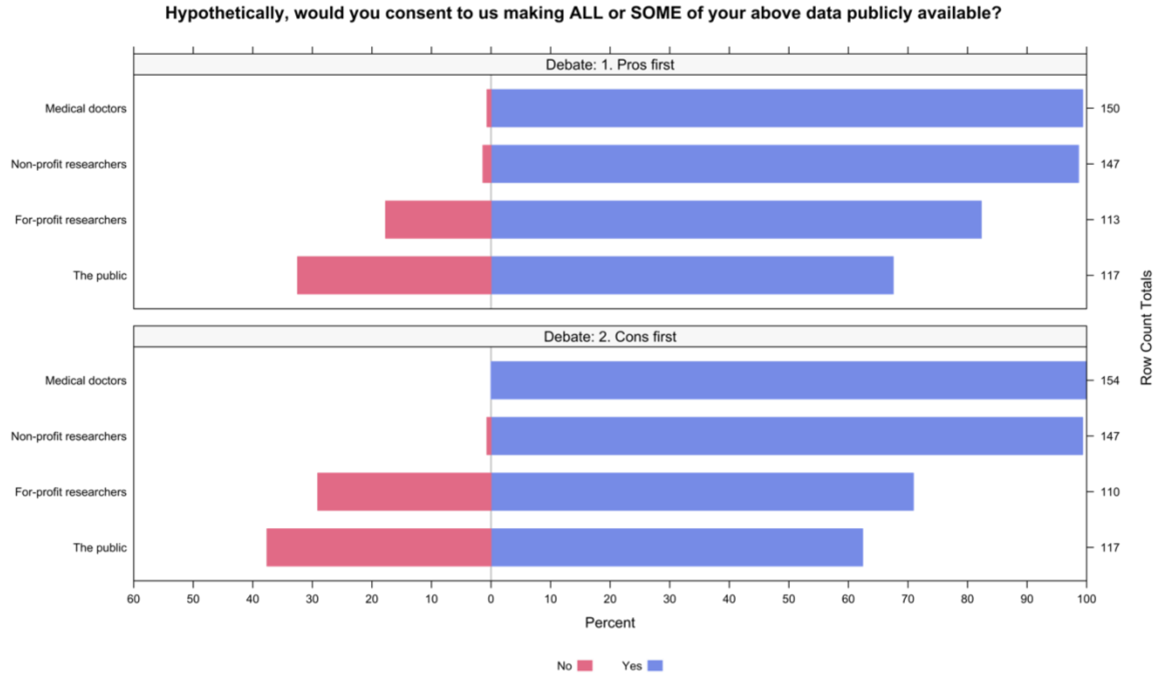

**Supplementary Figure 11. Respondents' views on researchers hypothetically sharing their treatment information by whether participants passed the comprehension check or not.**

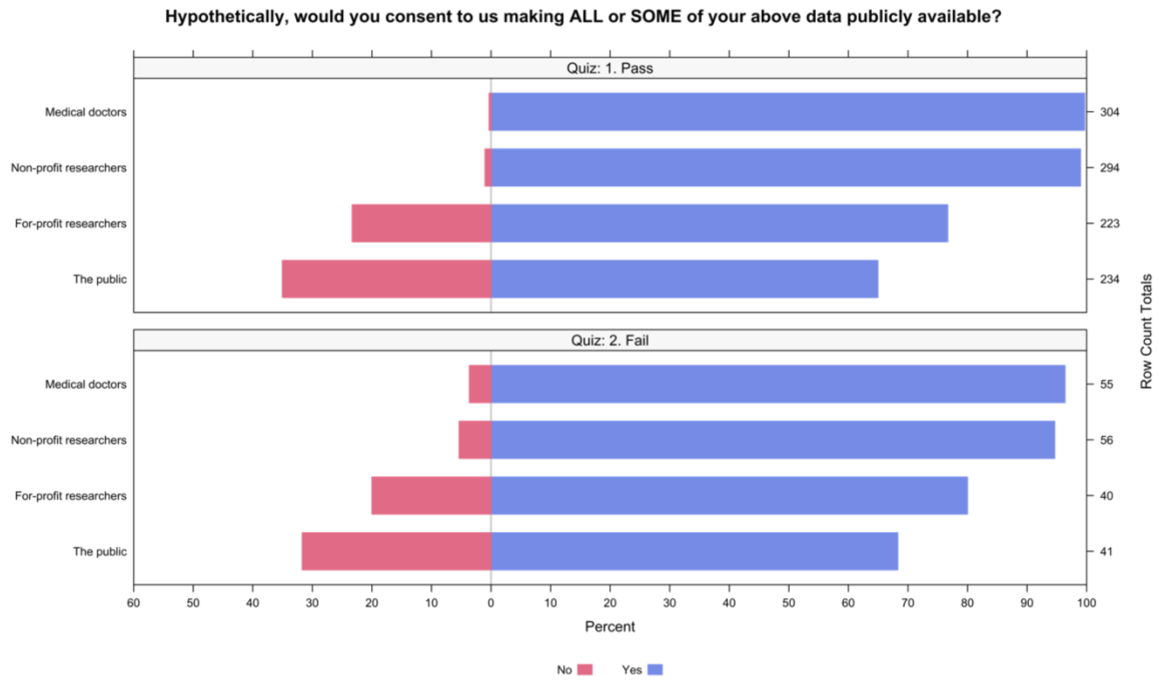

**Supplementary Figure 12. Respondents' views on the public sharing of the survey data by levels of trust in multiple stakeholders.**

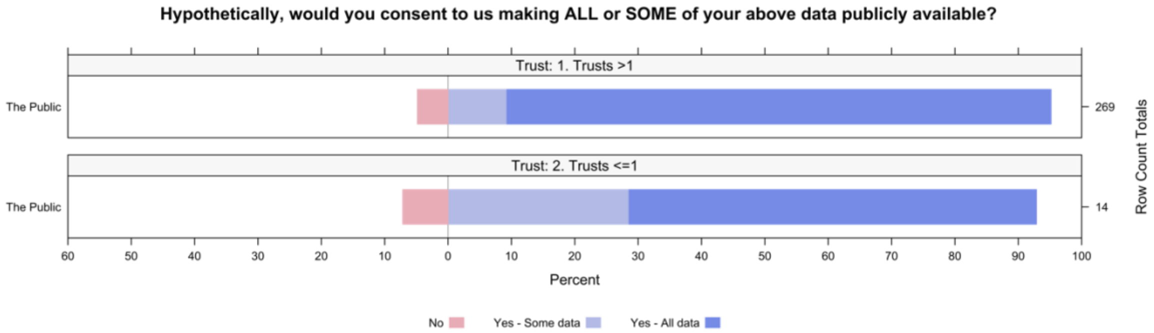


**Supplementary Figure 13. Respondents' views on the public sharing of the survey data by previous participation in research.**

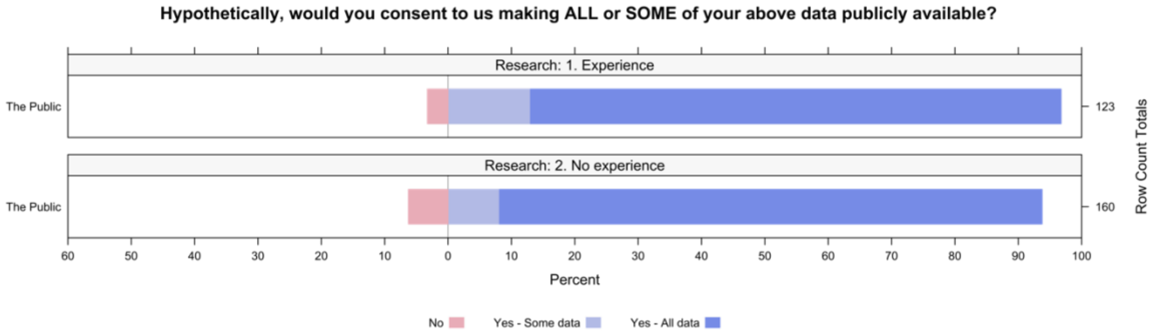


**Supplementary Figure 14. Respondents' views on the public sharing of the survey data by familiarity with data sharing.**

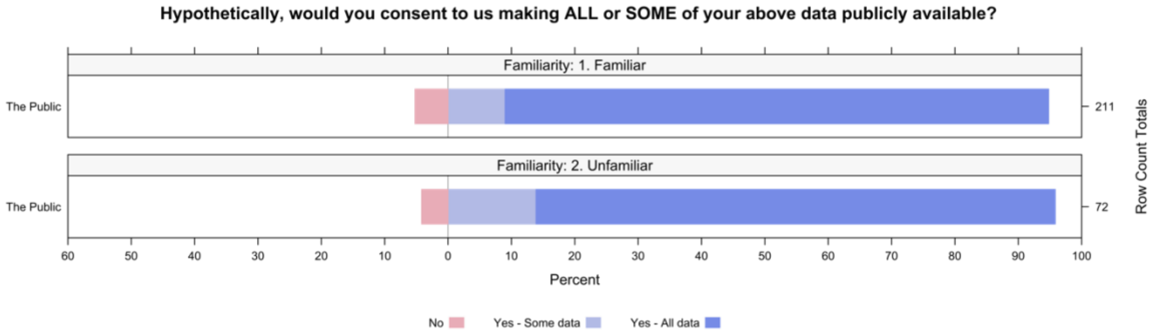

**Supplementary Figure 15. Respondents' views on the public sharing of the survey data by the order which participants were shown the advantages and disadvantages of sharing data.**

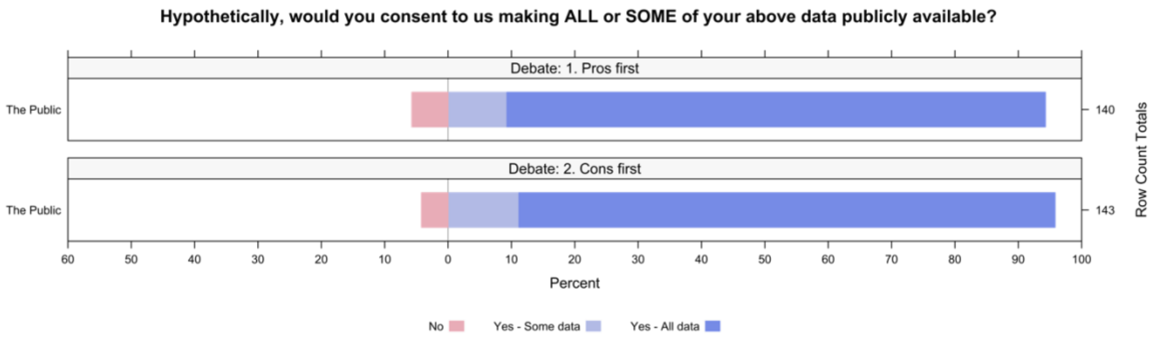


**Supplementary Figure 16. Respondents' views on the public sharing of the survey data by whether participants passed the comprehension check or not.**

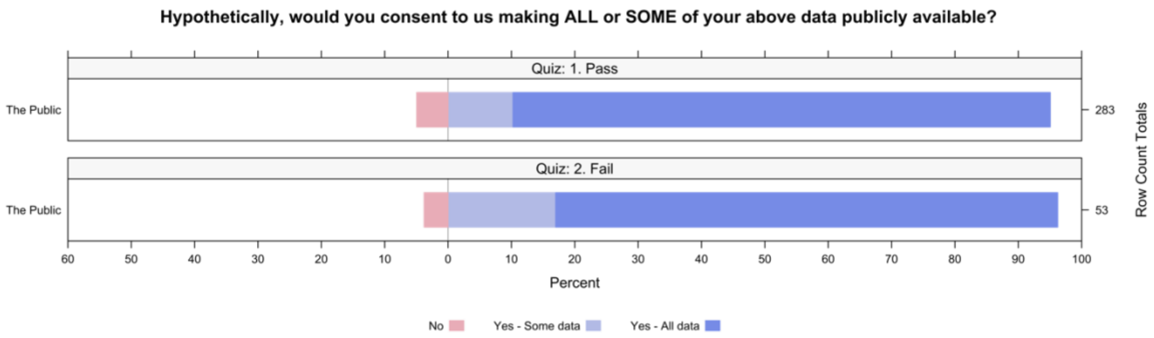
