## Appendix 2 for "What do Australians affected by cancer think about oncology researchers sharing research data: a cross-sectional survey"

### APPENDIX 2. SURVEY TRANSCRIPT

#### Block A: Consent *SMOG 7$^{th}$ Grade (11.32)*

**Please read the following information carefully before you decide to participate in the study.**

**What is this research about?** The aim of this study is to improve our understanding of what Australians diagnosed with cancer think about the sharing of data from medical research.

**What will I be asked to do?** You will be asked to complete a short anonymous survey. The survey should take less than 15 minutes to finish.

**Are there any risks?** The survey will ask you basic questions about your cancer journey (e.g., type of cancer and time since treatment). As such, there is a chance that recalling these events may cause some people distress.

**Are there any benefits?** There are no direct benefits for completing the survey. However, your answers may help change future research and practices on sharing research data.

**Do I have to take part?** Participation in this survey is completely optional. If you do start the survey, you can skip any question you do not want to answer or stop the survey whenever you want. However, whilst you can stop at any time, it will not be possible to remove any answers you give before stopping. This is because your responses are all anonymous and so we cannot identify which are yours for removal.

**What will happen to information about me?** All your answers to the survey will be stored indefinitely on a secure server hosted by a University of Melbourne approved survey provider (Qualtrics) and safeguarded subject to legal requirements. Survey responses will only be accessible by the named researchers. At the conclusion of the project, any potentially identifiable details will be removed, and the data will be stored privately on the Open Science Framework in perpetuity, and will be made available only on request to other researchers for future research.

**Will I hear about the results of this project?** The results of the survey will be reported in conference presentations and scientific articles. After you finish the survey you can join a private mailing list to be sent the results once they have been published.

**Where can I get further information?** If you would like more information about the project, please contact Professor Fiona Fidler, Responsible Researcher.

**Who can I contact if I have any concerns about the project?** This project has human research ethics approval from The University of Melbourne (Ethics ID: 22111). If you have any concerns or complaints about the conduct of this research project, which you do not wish to discuss with the research team, you should contact the Research Integrity Administrator, Office of Research Ethics and Integrity, University of Melbourne, VIC 3010. Tel: +61 8344 1376 or. All complaints will be treated confidentially. In any correspondence please provide the name of the research team and/or the name or ethics ID number of the research project.

---

**Consent**

If you would like to take part in this study, please indicate your agreement with the following statements:

1. I consent to participate in this project, the details of which have been explained to me, and I have been provided with a copy of the Plain Language Statement to keep.
2. I understand that the purpose of this research is to understand what Australians diagnosed with cancer think about the sharing of data from medical research.
3. I understand that my participation in this project is for research purposes only.
4. I acknowledge that the possible effects of participating in this research project have been explained to my satisfaction. In this project I will be asked to complete a short anonymous online survey.
5. I understand that my participation is voluntary and that I am free to stop the survey anytime without explanation or prejudice, but that any recorded responses cannot be withdrawn.
6. I understand that my answers to the survey will be stored indefinitely at the University of Melbourne.
7. I understand that anonymised data from this research will be stored privately on the Open Science Framework in perpetuity and will only be made accessible on request to other researchers for future research.
8. I have been informed that the information I provide will be safeguarded subject to any legal requirements.
9. I understand that by proceeding to and completing the survey I am signifying my consent for the data to be used in the above contexts.
10. I understand that after I complete this consent form, it will be retained by the researcher.

**By clicking the yes button below, I acknowledge that I have read the above statements, I am 18 years old or older, and I agree to participate:**

**Thank you for agreeing to participate in the survey. Before we begin, we want to ask you some questions about yourself.**

Please note: All answers to the survey are ANONYMOUS and will NOT be shared publicly. You also do not have to answer any questions you don't want to.

**Q1. Where did you hear about this study? (Select all that apply.)**

- ◯ Facebook
- ◯ Twitter
- ◯ LinkedIn
- ◯ Online Newsletter
- ◯ Flyer in hospital clinic
- ◯ Word of mouth
- ◯ Other, please specify:

**Q2. What is your age?**

_________

**Q3. What is your gender?**

- ◯ Male
- ◯ Female
- ◯ Prefer to self-describe as:

**Q4. Do you currently live in Australia?**

- ◯ Yes
- ◯ No

**Q5. Do you live in a …**

- ◯ Metropolitan area (population greater than 100,000 people)
- ◯ Small or large rural town (population between 10,000 to 100,000 people)
- ◯ Remote or very remote community (population less than 10,000 people)

**Q6. What is your highest level of education?**

▼ Postgraduate Degree ... Did not go to school

**Q7. What cancer were you diagnosed with?** (Note: If you have been diagnosed with multiple cancers please select the one that occurred most recently.)

▼ Anal cancer ... Other cancer

**Q8. How long has it been since your last treatment for the above cancer?**

- ◯ Currently receiving treatment
- ◯ Less than a year ago
- ◯ 1-5 years ago
- ◯ 6-10 years ago
- ◯ 10+ years

**Q9. Have you previously participated in a cancer-related research project? (Select all that apply.)**

- ◯ Yes - Clinical trial
- ◯ Yes - Other research
- ◯ No
- ◯ I'm not sure

**Q10. Would you generally trust the following people with your personal information?**

| | I would generally NOT trust | I'm not sure | I would generally trust |
|---|---|---|---|
| a) My General Practitioner (GP) | ◯ | ◯ | ◯ |
| b) Other medical doctors in Australia | ◯ | ◯ | ◯ |
| c) Researchers at an Australian university | ◯ | ◯ | ◯ |
| d) Researchers at an Australian company | ◯ | ◯ | ◯ |
| e) The Australian government | ◯ | ◯ | ◯ |

**For this survey we are interested in getting your views on the sharing of research data. But when we say ‘research data’ what do we mean?**

Cancer scientists do research for many reasons. They might want to test how well a new treatment works, improve ways of finding cancers earlier, or better understand how cancers grow.

When scientists run experiments, they will record information about the methods they use, the subjects of the research (people, tissues, cells) and the results of each test or observation. This raw information is normally referred to as ‘**research data**’.

When experiments are completed, cancer scientists will make a **summary** of their **research data** public to share discoveries, improve practices and inform future research.

Below is an example of what research data (green text) and a summary (yellow text) might both look like.

| | Case | Age | Sex | Weight | Drug | Side effects |
|---|---|---|---|---|---|---|
| **RESEARCH DATA** | A | 60 | Female | 73 kg | 1 | Yes |
| | B | 35 | Male | 88 kg | 2 | No |
| | C | 90 | Male | 63 kg | 2 | No |
| | D | 54 | Male | 103 kg | 1 | Yes |
| | E | 21 | Female | 68 kg | 1 | Yes |
| | ↓ | ↓ | ↓ | ↓ | ↓ | ↓ |
| **SUMMARY** | 5 people were studied | The average age was 52 years | 40% were female | The average person weighed 79 kg | 60% received Drug 1 | All people on Drug 1 experienced side effects |

While cancer scientists will always share a **summary** of their research data publicly, it is not yet standard practice to share **research data** publicly as well. (Please note: when sharing any information collected from people, scientists will ALWAYS remove important personal information (e.g., name, date of birth) to prevent anyone finding out who the participants were i.e., 'anonymise' or 'de-identify' the data.)

**This survey plans to ask you what you think about cancer scientists sharing research data. For this survey, at times we want you to imagine that you might be Case A, B, C, D or E.**

**Why do cancer researchers often not share their research data?**

There is a debate among scientists about whether sharing research data is a good or bad idea. Some of the reasons for and against sharing include the following.

Reasons to share.
- It can speed up discoveries by allowing information from multiple studies to be combined.
- It can allow others to answer new questions not thought of by the original researchers.
- It can allow others to check for mistakes.
- It can allow others to calculate other useful results.

Reasons NOT to share.
- Some scientists are afraid other researchers may misuse their data.
- Other researchers may make discoveries using shared data before the scientists who collected it have a chance to.
- There may be a risk of somebody figuring out who research participants were (e.g., patients with very rare cancers).
- It can be difficult to share depending on who owns the research data and how big the files are.

**Just to check this all makes sense we have a quick quiz for you.**

The following two pieces of information are from a 2019 study of 12 patients with lymphoma who received treatment for viral and fungal infections.

**INFORMATION #1.**

> "Among the 12 cases of infection, the average age was 66 years (range 34-78) and the male/female ratio was 1:1. … The infection rate at 28 days indicated a poor prognosis (infection rate of 50%; 6/12)."

**INFORMATION #2.**

| Case | Age | Sex | Infection | Outcome |
|---|---|---|---|---|
| 1 | 34 | Male | Fungal | Not improved |
| 2 | 71 | Female | Fungal | Not improved |
| 3 | 65 | Male | Fungal | Improved |
| 4 | 73 | Female | Fungal | Not improved |
| 5 | 74 | Female | Fungal | Improved |
| 6 | 56 | Female | Viral | Improved |
| 7 | 67 | Male | Viral | Improved |
| 8 | 78 | Female | Viral | Not improved |
| 9 | 74 | Male | Viral | Not improved |
| 10 | 72 | Female | Viral | Not improved |
| 11 | 54 | Male | Fungal | Improved |
| 12 | 78 | Male | Fungal | Improved |

**Q11. Please select which you think is the research data and which is the summary.**

- ◯ #1 is the **Research Data** and #2 is the **Summary**. [Incorrect answer]
- ◯ #1 is the **Summary,** and #2 is the **Research Data**. [Correct answer]
- ◯ Neither of the above is correct. [Incorrect answer]

**Q12. Are you familiar with what has been described in the previous sections? (i.e., what research data is, the debate about the reasons for and against sharing.)**

- ○ Never heard about this before. [Proceed to Q14]
- ○ I've heard some of these things but not sure what they mean. [Proceed to Q13]
- ○ I know a little about this topic [Proceed to Q13]
- ○ I know a lot about this topic. [Proceed to Q13]
- ○ I'm an expert on data sharing. [Proceed to Q13]

**Q13. Can you tell us why you are familiar with these topics? (Select all that apply.)**

- ○ I'm a health professional that primarily cares for cancer patients (e.g., oncologist, specialist oncology nurse, radiotherapist).
- ○ I'm a health professional (e.g., nurse, GP, surgeon, hospital specialist, allied health professional, hospital administration staff).
- ○ I'm a health researcher/student.
- ○ I'm a researcher/student in a field outside of health.
- ○ I have participated in health research previously where this was explained to me.
- ○ I'm interested in how scientific research is conducted.
- ○ Other, please specify:

**Based on your understanding of what 'research data' is, and assuming researchers are safely and legally able to do so:**

**Q14. Do you think cancer researchers should regularly share data collected from <u>NON-HUMAN research participants</u> (e.g., animals, cells, viruses) with…?**

| | Feel STRONGLY that researchers should NOT share | Feel that researchers should NOT share | Indifferent | Feel that researchers should share | Feel STRONGLY that researchers should share |
|---|---|---|---|---|---|
| Medical doctors (e.g., to improve treatments and also get scientific publications). | ○ | ○ | ○ | ○ | ○ |
| Non-profit researchers (e.g., to do medical research and also bring in new funding). | ○ | ○ | ○ | ○ | ○ |
| For-profit researchers (e.g., to develop medicines and also make money for shareholders). | ○ | ○ | ○ | ○ | ○ |
| The public (e.g., deposit the data into a publicly accessible repository to allow any interested researcher to access and use it for any purpose). | ○ | ○ | ○ | ○ | ○ |

**Q15. Do you think cancer researchers should regularly share ANONYMOUS data collected from HUMAN participants with…?**

| | Feel STRONGLY that researchers should NOT share | Feel that researchers should NOT share | Indifferent | Feel that researchers should share | Feel STRONGLY that researchers should share |
|---|---|---|---|---|---|
| Medical doctors (e.g., to improve treatments and also get scientific publications). | ○ | ○ | ○ | ○ | ○ |
| Non-profit researchers (e.g., to do medical research and also bring in new funding). | ○ | ○ | ○ | ○ | ○ |
| For-profit researchers (e.g., to develop medicines and also make money for shareholders). | ○ | ○ | ○ | ○ | ○ |
| The public (e.g., deposit the data into a publicly accessible repository to allow any interested researcher to access and use it for any purpose). | ○ | ○ | ○ | ○ | ○ |

**Q16. What percentage of cancer research do you think currently shares research data?**

0 10 20 30 40 50 60 70 80 90 100

Guess (%)

**For this question we want you to imagine that some of your medical information stored at the location where you received treatment for your diagnosed cancer was going to be collected for a research project.**

Note: When thinking about possible information collected for cancer research, we want you to think of information that could be stored in a person's medical records. For example: weight and blood pressure measurements, blood test results, through to disease history and prescribed treatments.

**Q17. If the researchers asked you, would you consent to the researchers sharing ANONYMOUS research data with…?**

| | Yes | No | Unsure |
|---|---|---|---|
| Medical doctors (e.g., to improve treatments and also get scientific publications). | ○ | ○ | ○ |
| Non-profit researchers (e.g., to do medical research and also bring in new funding). | ○ | ○ | ○ |
| For-profit researchers (e.g., to develop medicines and also make money for shareholders). | ○ | ○ | ○ |
| The public (e.g., deposit the data into a publicly accessible repository to allow any interested researcher to access and use it for any purpose). | ○ | ○ | ○ |

**Here is a picture of what the research data from THIS SURVEY might look like (including some of your responses so far in yellow).**

| | PARTICIPANT INFORMATION | | | | | | | | | SURVEY | |
|---|---|---|---|---|---|---|---|---|---|---|---|
| Case | Age | Gender | Australian resident | Rurality | Education | Cancer | Time since treatment | Research participant | Trust | Quiz | Familiar |
| A | 23 | Female | Yes | Metropolitan | Bachelor Degree | Breast cancer | Ongoing | Yes - Clinical trial | 1,1,1,1,1 | 1 | 1 |
| B | 39 | Male | Yes | Metropolitan | High School | Lung cancer | <1 year | No | 1,1,1,1,1 | 1 | 0 |
| C | 35 | Male | Yes | Metropolitan | Postgraduate Degree | Prostate cancer | Less than a year ago | Yes - Clinical trial | 1,1,1,0,1 | 1 | 4 |
| D | 80 | Female | Yes | Rural | Did not go to school | Breast cancer | 10+ years | No | 1,1,1,1,1 | 1 | 0 |
| E | 64 | Male | Yes | Metropolitan | High School | Prostate cancer | 5-10 years | Yes - Other | 1,1,0,0,1 | 1 | 0 |
| F | 55 | Female | Yes | Metropolitan | Postgraduate Degree | Skin cancer | <1 year | No | 1,0,0,0,0 | 1 | 2 |

**Q18. Once the summary of THIS SURVEY has been made public do you think the research data should be permanently deleted?**

- ◯ Yes.
- ◯ No.
- ◯ I'm not sure.

**Q19. Once the summary of THIS SURVEY has been made public do you think it's appropriate for us (the research team) to keep the research data for an indefinite period of time?**

- ○ Yes.
- ○ No.
- ○ I'm not sure.

**Reminder: The data from THIS SURVEY will NOT be shared publicly.**

**Q20. Hypothetically, would you consent to us (the research team) making ALL or SOME of your above data publicly available?** (e.g., deposit the above ANONYMISED data into a freely accessible repository to allow any interested researcher to access and use it for any research purpose.)

- ○ Yes - All data. [Proceed to Q23]
- ○ Yes - Some data. [Proceed to Q21]
- ○ No. [Proceed to Q22]
- ○ I'm not sure. [Proceed to Q22]

**Here is a picture of what the research data from THIS SURVEY might look like (including some of your responses so far in yellow).**

| | PARTICIPANT INFORMATION | | | | | | | | | SURVEY | |
|---|---|---|---|---|---|---|---|---|---|---|---|
| | A | B | C | D | E | F | G | H | I | J | K |
| Case | Age | Gender | Australian resident | Rurality | Education | Cancer | Time since treatment | Research participant | Trust | Quiz | Familiar |
| A | 23 | Female | Yes | Metropolitan | Bachelor Degree | Breast cancer | Ongoing | Yes - Clinical trial | 1,1,1,1,1 | 1 | 1 |
| B | 39 | Male | Yes | Metropolitan | High School | Lung cancer | <1 year | No | 1,1,1,1,1 | 1 | 0 |
| C | 35 | Male | Yes | Metropolitan | Postgraduate Degree | Prostate cancer | Less than a year ago | Yes - Clinical trial | 1,1,1,0,1 | 1 | 4 |
| D | 80 | Female | Yes | Rural | Did not go to school | Breast cancer | 10+ years | No | 1,1,1,1,1 | 1 | 0 |
| E | 64 | Male | Yes | Metropolitan | High School | Prostate cancer | 5-10 years | Yes - Other | 1,1,0,0,1 | 1 | 0 |
| F | 55 | Female | Yes | Metropolitan | Postgraduate Degree | Skin cancer | <1 year | No | 1,0,0,0,0 | 1 | 2 |

**Q21. Which of the above information, if any, would you NOT want to be shared publicly? (Select all columns that apply.)**

- ◯ Column A - Age.
- ◯ Column B - Gender.
- ◯ Column C - Australian resident.
- ◯ Column D - Rurality.
- ◯ Column E - Education.
- ◯ Column F - Cancer diagnosis.
- ◯ Column G - Time since treatment.
- ◯ Column H - Previous research participant status.
- ◯ Column I - Trust.
- ◯ Column J - Quiz results.
- ◯ Column K - Familiarity with data sharing.
- ◯ Would consent to sharing all of the data.

**Q22. What concerns would you have about the public sharing of the data from THIS SURVEY?**

________________________________________________________________

________________________________________________________________

________________________________________________________________

**Q23. Do you have any other thoughts about data sharing or the survey which you'd like to express?**

________________________________________________________________

________________________________________________________________

________________________________________________________________

**That's it for the main survey. However, if you are happy to keep going, we do have two more bonus questions we would like to ask you, which you can see by clicking the next button. If you'd prefer to stop, we thank you very much for your participation. Your responses have been recorded and you can now close your browser window.**

Please also note, if you would like to be informed about the results of this research you can follow this link to join the study's mailing list.

We would also greatly appreciate if you could share the link to this survey with any of your family members, friends and colleagues who have been affected by cancer and would be interested in adding their voice to this topic.

*Furthermore, if completing this survey has also brought up difficult emotions for you, please reach out to your general practitioner, community nurse or call the Cancer Council Helpline on 13 11 20 for information, resources and connection to support programs and advocacy services for those affected by cancer. If you are in immediate danger, please call 000.*

**BONUS ROUND:**

In medicine the results of clinical trials are considered one of the most trustworthy forms of scientific evidence that can be generated.

As a consequence, the medical community relies on researchers running trials to publish timely summaries of their results in order to share new discoveries, identify areas for future research (as well as research 'dead ends'), and ultimately to ensure patients receive the best and safest medical treatments available.

However, previous research has shown that approximately half of clinical trials fail to report their results in a timely manner, or at all.

For example, one of these studies reported that between 2006 and 2016, over 11,000 completed clinical trials (enrolling 8.7 million patients) still had not shared their results with doctors, researchers, or patients.

Appreciating this, we would like to ask you the following questions:

**BONUS QUESTION 1: Do you think what has been described above would be important information for people to know when considering participation in a clinical trial?**

- ○ Not important at all.
- ○ Somewhat important.
- ○ Very important.
- ○ Absolutely essential.

**BONUS QUESTION 2: Would finding out that the cancer research you previously participated in never shared its results make you more or less likely to participate in another project from the same research team in the future?**

- ○ Much less likely to participate.
- ○ Slightly less likely to participate.
- ○ Would not influence my decision to participate.
- ○ Slightly more likely to participate.
- ○ Much more likely to participate.

**BONUS QUESTION 3: Would being told that there was a 50% chance that the results would not be shared make you more or less likely to participate in a clinical trial in the future?**

- ○ Much less likely to participate.
- ○ Slightly less likely to participate.
- ○ Would not influence my decision to participate.
- ○ Slightly more likely to participate.
- ○ Much more likely to participate.
